# A scoping umbrella review to identify anti-racist interventions to reduce ethnic disparities in health and care

**DOI:** 10.1101/2023.05.03.23289263

**Authors:** Jennifer L Y Yip, Shoba Poduval, Leah de Souza-Thomas, Sophie Carter, Kevin Fenton

**Affiliations:** Consultant Lead for Health Equity, Science and Strategy, Office for Health Improvement and Disparities, London Region, Department of Health and Social Care, 39 Victoria Street, London SW1H 0EU, United Kingdom; Academic Clinical Lecturer in Primary Care & Population Health, Research Department of Primary Care & Population Health, University College London, Royal Free Campus, Rowland Hill Street, London NW3 2PF, United Kingdom; Health Improvement Manager, Office for Health Improvement and Disparities, London Region, Department of Health and Social Care, 39 Victoria Street, London SW1H 0EU, United Kingdom; Project Manager, Health Innovation Manchester, Suite C, Third Floor, Citylabs, Nelson Street, Manchester, M13 9NQ, United Kingdom; Regional Director Office for Health Improvement and Disparities, London Region, Department of Health and Social Care, 39 Victoria Street, London SW1H 0EU, United Kingdom, Regional Director of Public Health NHS London, Statutory Health Advisor to the Mayor of London, GLA and London Assembly

## Abstract

**Objectives:** To identify anti-racist interventions which aim to reduce ethnic disparities in health and care.

**Eligibility criteria for selecting studies:** Only studies reporting systematic reviews of anti-racist interventions were included. Studies were excluded if no interventions were reported, no comparators reported, or the paper was primarily descriptive.

The following databases were searched: Embase, Medline, Social Policy and Practice, Social care online and Web of Science. Quality appraisal (including risk of bias) was assessed using the AMSTAR-2 tool.

Due to the nature of the selected reviews, the lack of meta-analyses and heterogeneity of included studies, a narrative synthesis using an inductive thematic analysis approach was conducted to integrate and categorise the evidence on anti-racist interventions for health and care.

**Results:** A total of 18 systematic reviews are included in the final review. 15 are from the healthcare sector and three are from education and criminal justice. 17 reviews are focused on interventions and one focused on implementation. All 18 reviews described interventions which targeted individuals and their communities, and 11 reviews described interventions targeting both individuals and communities, and healthcare organisations. On an individual level, the most promising interventions reviewed include group-based health education led by professional staff and providing culturally tailored or adapted interventions. On a community level, participation in all aspects of care pathway development that empowers ethnic minority groups may provide an effective approach to reducing ethnic health disparities. Targeted interventions to improve clinician patient interactions and quality of care for conditions with disproportionately worse outcomes in ethnic minority groups show promise.

**Discussion:** Many of the included studies were low or critically low quality due to methodological or reporting limitations. The heterogeneity of intervention approaches, study designs, and limited reporting of cultural adaptation, implementation and lack of comparison with White ethnic groups limited our understanding of the impact on ethnic health inequalities.

In summary, for programme delivery, different types of pathway integration and providing a more person-centred approach with fewer steps for patients to navigate can contribute to reducing disparities. For organisations, there is an overemphasis on patient education and individual behaviour change rather than organisational change, and recommendations should include a shift in focus and resources to policies and practices that seek to dismantle institutional and systemic racism through a multi-level approach.

## Introduction

Health inequalities have increased since 2010, with an extending 10-year gap in life expectancy between people living in the most and least deprived areas of England. ^1^ The COVID-19 pandemic augmented pre-existing inequalities and highlighted the impact of our social environment on our health. Rudolf Virchow was one of the first physicians to identify medicine as a social science. ^2^ Since then, health inequalities and the importance of social causes of poor health have been highlighted in UK public health policy by the Black report, ^3^ the Acheson report^4^ and the WHO Commission on Social Determinants of Health^5^. People living in poorer areas, and from less affluent backgrounds, have a higher risk of morbidity and mortality. ^6^ These conditions of human existence have existed for centuries, etched in science and literature. The interplay between social and economic factors also drives racial health inequalities, where communities from minoritised ethnic groups who live in areas of greater socioeconomic disadvantage, also experience additional drivers such as racism. ^7^

Since the start of the COVID-19 pandemic, the association between ethnicity and adverse health outcomes have risen in prominence. ^8–11^ People from Black, Asian and minority ethnic backgrounds were exposed to higher risk of morbidity and mortality from SARS-CoV-2, with the south Asian and Black populations suffering up to 5 times the risk of Covid deaths compared to their White counterparts.^12^ Differential access to interventions such as vaccinations and well-fitting face masks compounded the increased risk. ^13 14^ Though racial health disparities are not new; scientific awareness was coupled with cultural change that arose with the murder of George Floyd and Black Lives Matters movement. The National Health Service (NHS) Race and Health Observatory (RHO) was commissioned, and there are emerging efforts to address racial disparities in health and healthcare. There is extensive literature that demonstrates the association between minoritised ethnicity and poorer health outcomes. ^7 15^ In the UK, Black women are 4 times more likely to die in childbirth compared to White mothers, and experience around 4 times risk of stillbirths even after accounting for area deprivation and maternal age. ^16 17^ In addition to maternal ethnic health disparities, the NHS RHO also reported barriers to access in mental health and other aspects of care for ethnic minority groups that contribute to worse outcomes. ^18^

Racism is a driver of ethnic health inequalities, operating directly through discrimination and stigma, and indirectly through the social determinants of health. ^7 19^ The social patterns that mediate health inequalities such as differential access to material, social and healthcare resources, health behaviours, psychosocial stress, also reflects racialised patterns and highlights the intersectional nature of health risk. The focus on social inequalities in UK health policy has to some extent, masked the impact of race and racism on health disparities because racism drives, and its effects are mediated through structured social and economic inequalities. ^19^ Abubakar and colleagues reviewed a wide range of literature that examined the relationship between racism, xenophobia, discrimination and health outcomes, and outlined key areas for intervention from the global literature. ^20^ Further examination is required to implement and recommend actions from this wide-ranging approach. In the UK, particularly in diverse urban areas, there is an urgent need to take action to mitigate the impact of racism on adverse health outcomes.

The purpose of this review is to provide evidence to inform practice rather than to answer a specific clinically meaningful question, and therefore a scoping review with a narrative synthesis was chosen as the most appropriate approach. ^21^ We sought to identify which anti-racist interventions are effective in the health and care sector, focusing our synthesis on potential implementation in the UK health and care system.

### Definitions

For the purpose of this study, we reviewed a number of commonly used definitions of racism and anti-racism to inform the selection of studies. A full discussion on the definitions is beyond the scope of this review. In this study, we draw on Ibrahim X. Kendi’s definition of anti-racism as “a powerful collection of antiracist policies that lead to racial equity and are substantiated by antiracist ideas”, where an antiracist idea is “any idea that suggests the racial groups are equals in all their apparent differences.”^22^ The focus of anti-racist work in the health and care sector is therefore to reduce racial health inequalities, which also resonates with Powell’s definition of anti-racism ^23^, and the aim of anti-racist work in health and care is working towards racial health equity.

We also recognise the difference in definitions between race and ethnicity, but here we will use the terms interchangeably.

An important consideration we identified from different definitions of racism is the role of power dynamics.^23–28^ Therefore, in our selection and definition of anti-racist interventions the role of power differentials was an important consideration.

One commonly used definition of anti-racist intervention is from Calliste and Dei: an “action-oriented, educational and/or political strategy for systemic and political change that addresses issues of racism and interlocking systems of social oppression”. ^29^ This definition addresses a wide range of actions in a number of settings, and at its core, recognises the systemic and embedded impact of racism and oppression. The role of people (agents) and structures and their power to emancipate or oppress has also been recognised in realist social theory conceptualised by Farr and Archer.^30^

In order to detect a “strategy for systemic change”, we sought to identify interventions that aimed to reduce ethnic health disparities, recognising that racism is embedded in the structures and processes which cause ethnic disparities, but may not be explicitly recognised in causal pathways. ^26 27^ Therefore we included studies that either define the intervention as anti-racist, and in those that did not, we included the interventions providing there was both

1. Evidence that the intervention redressed the imbalance of power. We used Farr’s implementation of Archer’s realist social theory framework to question the role of structures and agents and their potential impact on power dynamics in the description of the interventions.^30^
2. Explicit ambition to reduce ethnic or racial health disparities

### Framework

To categorise the different interventions identified, we used an adaptation of Dahlgren and Whitehead’s socio-ecological model showing racism as a driver of different levels of social determinants of health. *(Figure 1.)* Here, we show structural racism acting at the highest level of society; and implied in the diagram is the impact of the wider determinants on more proximal risk factors of disease resulting in ethnic health inequalities, as racism is embedded and causes harm whilst hidden in our health and care system, institutions, policies, cultures and behaviours, growing over time.

**Figure 1.**
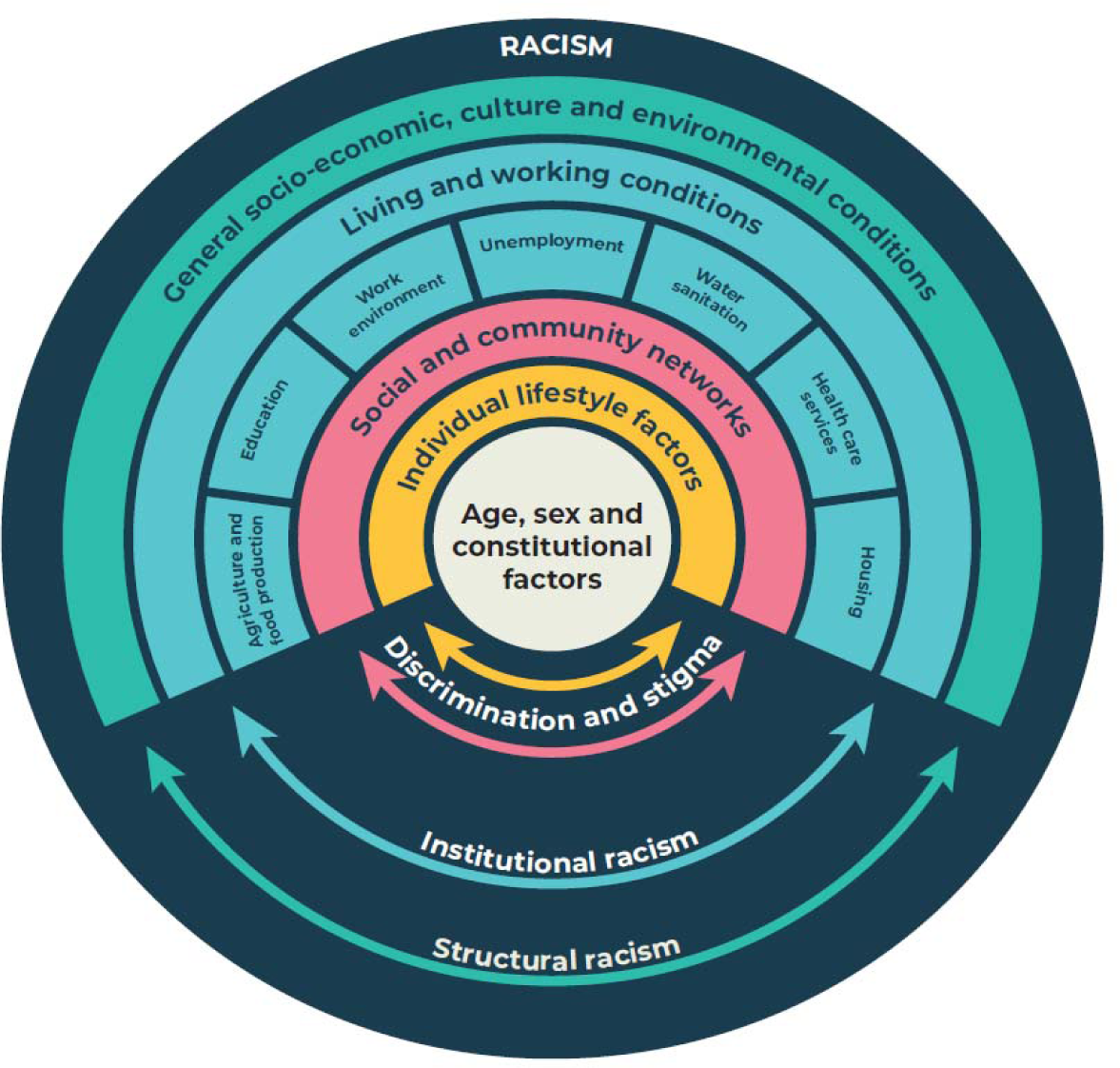
Conceptualising racism integrated with the social determinants of health. ^31^ Adapted from Dahlgren and Whitehead, 1993, showing racism as a driving force for social determinants of health. Though social determinants are universal, racism is one of a range of driving forces that exists in our societies and that acts on these determinants. ^32^

## Methods

We conducted a scoping literature review in accordance with recognised methodology ^21^ and reported in line with the Preferred Reporting for Systematic reviews and Meta-Analyses (PRISMA) statement.^33^ The search strategy, inclusion and exclusion criteria, appraisal tool and data collection instruments were designed and agreed prior to selection. We made one change from protocol to formally exclude reviews from the educational literature after data extraction.

### Search strategy

A literature search of the following databases was conducted in collaboration with a knowledge and evidence specialist: Embase, Medline, Social Policy and Practice, Social care online and web of science, with the aim to identify “what works in antiracism” focusing on interventions and programmes that addressed racism. We included search terms based on Medical Subject Headings (MeSH) and keywords related to race, ethnicity, racism and anti-racism. A full list of the search terms and the search strategy is included in the Appendix 1.

### Eligibility criteria

We limited the search to papers published from the year 2000 onwards as initial searches indicated that more literature emerged after this date.

Inclusion criteria were:

1. Studies reporting systematic reviews only, where it was stated in the title or methods and, with explicit inclusion and exclusion criteria and where authors searched more than one literature database;
2. Interventions were anti-racist, based on definitions above. We sought evidence that the intervention redressed the imbalance of power by asking the question: “Do the interventions address differences in power and reallocate resources to people from minoritised ethnic groups?”
3. Publication was available in English language

Exclusion criteria were:

1. No interventions reported
2. No comparators reported and the paper was primarily descriptive, for example a review of epidemiological studies of association between race and an outcome.

#### Selection process

The selection process is outlined in *Figure 2.* using the PRISMA 2020 statement on guidelines for reporting systematic reviews. One of the first authors (JY) drafted the protocol and data collection table, and together with two co-authors (SC and LDT) agreed with the final protocol prior to conducting data collection. Two authors (SC and JY) independently reviewed titles and abstracts for inclusion, with consensus in discussion with third author (LDT). Data extraction was conducted independently by three authors (SC, JY and SP) using a data extraction form based on the study objectives. In our selection process, we found two reviews looking at educational interventions, ^34 35^ which we considered as a separate category and did not formally include in our analysis and synthesis. Our search criteria only included databases that focused on health and social care, though our search terms included educational interventions and one looking at criminal justice. ^34–36^ which we considered as a separate category in our analysis and synthesis. Our search criteria only included databases that focused on health and social care, though our search terms included educational interventions.

**Figure 2.**
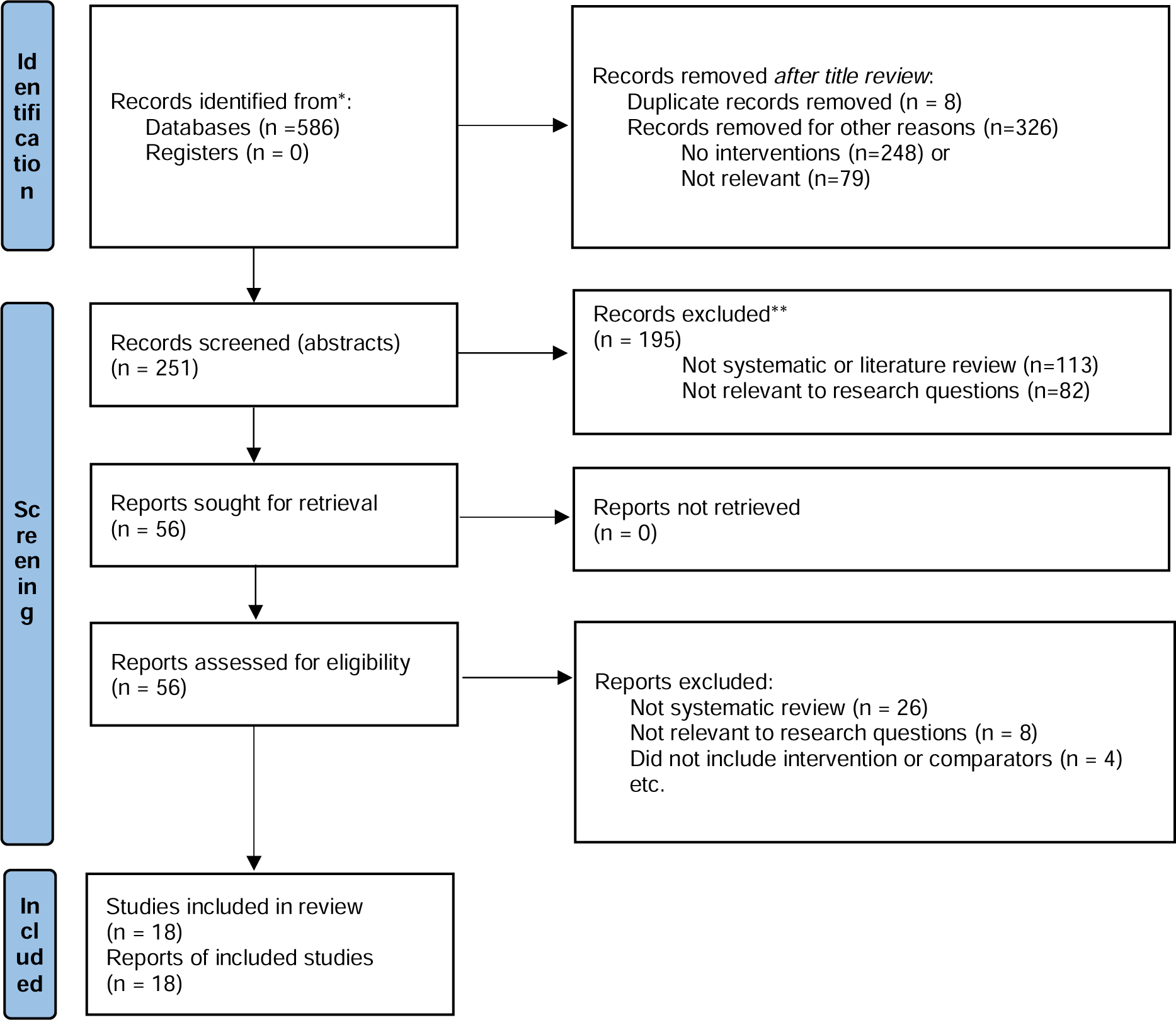
PRISMA diagram of study selection process

#### Quality assessment/appraisal (tools)

Two authors (JY and SP) independently used the AMSTAR-2 tool to evaluate quality of the selected systematic reviews.^37^ This resulted in a qualitative assessment of the studies, and we took the quality into account in the synthesis of the evidence. We prioritised intervention reported in reviews that were considered higher quality, or where there was consistent evidence across several reviews.

### Data synthesis

Framed by the socio-ecological and racism model outlined above, we used an inductive thematic analysis approach combined with an effectiveness review to integrate and categorise the evidence on anti-racist interventions for the health and care system. Due to the nature of the selected reviews, the lack of meta-analyses and heterogeneity of included studies, we conducted a narrative synthesis.

### Patient and public involvement

Due to the nature of the study, with secondary analysis of systematic review; there was no direct patient involvement in the conception or conduct of the study.

## Results

A total of 586 records were reviewed at the identification stage, eight were removed due to duplication and an initial title screen resulted in 251 records where abstracts were reviewed. A total of 56 studies were selected for retrieval and 18 reviews were included in the final selection.

Of the 18 systematic reviews included in the final review (see Figure 2), 15 were from the healthcare sector and three from outside of healthcare (education and criminal justice). 17 reviews are focused on interventions and one focused on implementation.

Here, we describe an intervention that either has an anti-racist focus, or interventions that address differences in power and reallocate resources to people from ethnic minority groups to produce race equity.

### Description of included studies

A total of 816 individual studies were included across all systematic reviews. Ten reviews specified health conditions in their inclusion criteria (cardiovascular disease ^38^, cancer screening ^39 40^, mental health ^41 42^, palliative care ^43 44^, HIV ^45^, childhood obesity ^46^ and diabetes ^47^), five reviews did not specify and explored health disparities in general ^48–53^, and three did not focus on health conditions ^34 36 54^. Ten reviews included any ethnic minority (non-white) group ^38–40 42 43 46–49 51 53^, four reviews focused on one ethnic group (black, African, African diaspora or Indigenous peoples ^36 41 44 45^), and four reviews did not specify ethnic criteria ^34 50 52 54^. All 18 reviews described interventions which targeted individuals and their communities, and 11 reviews described interventions targeting both individuals and communities, and healthcare organisations. A summary of characteristics of all included studies is available in table 1, and summary of interventions, outcomes and findings is outlined in Table 2.

**Table 1:** Summary of review characteristics (n=18)

| Review characteristics | Count (%) |
| --- | --- |
| **Study setting** |  |
| Community-based | 1 (5) |
| Healthcare | 14 (78) |
| Education | 2 (11) |
| Criminal justice | 1 (5) |
| **Health conditions** |  |
| Common chronic conditions (cardiovascular disease, diabetes, obesity) | 3 (16) |
| Cancer screening | 2 (11) |
| Palliative care | 2 (11) |
| Mental health | 2 (11) |
| Noncommunicable disease (HIV) | 1 (5) |
| Health condition not specified | 5 (28) |
| Did not focus on health | 3 (16) |
| **Target population** |  |
| Black/African/African diaspora | 3 (16) |
| Indigenous peoples | 1 (5) |
| Any ethnic minority groups | 10 (56) |
| Ethnicity not specified | 4 (21) |
| **Level of intervention** |  |
| Individual and community | 18 (100) |
| Healthcare organisation | 11 (61) |

**Table 2.** Summary of included studies

| Full Citation | Anti-racism intervention | Outcome measures | Main Findings |
| --- | --- | --- | --- |
| 1.Anderson LM, Adeney KL, Shinn C, Safranek S, Buckner-Brown J, Krause LK. Community coalition-driven interventions to reduce health disparities among racial and ethnic minority populations. Cochrane Database of Systematic Reviews 2015, Issue 6. Art. No.: CD009905. DOI: 10.1002/14651858.CD009905.pub2. Accessed 10 January 2023. | Locally recruited coalitions (groups of representatives) of racial and ethnic minority communities in partnership with social and health service agencies, schools, businesses, etc including 1) Community-based programs and policies ; 2) Health and social care system-level strategies that target neighbourhood social conditions influencing health outcomes (e.g. access to healthy food, safe neighbourhood environments) ; 3) Lay community health workers and group based health education targeting community risk behaviours (e.g. smoking). | Primary outcomes are direct measures of health status and lifestyle factors, mortality (e.g. all-cause death within period of study, probability of survival), morbidity (e.g. quality of life measures, incidence rates, measures of symptoms and functionality) and health behaviour change measures (e.g. measures of physical activity, smoking status, alcohol consumption, dietary change). Secondary outcomes are changes in neighbourhood conditions or policies introduced to promote community health improvement (e.g. a policy establishing a farmers' market to provide access to fresh produce, a school policy opening sports fields for local resident use during non-school periods). | Lay community health outreach worker interventions (n = 10) and group-based health education led by professional staff (n = 5) produced fairly consistent positive effects on health and behavioural outcomes. There were mixed results from peer-based education on behavioural outcomes, including improved BMI from church-based peer interventions in two studies. These findings provide evidence that community coalition-driven interventions can benefit minority populations. Findings are limited by general conclusions from a wide range of interventions targeting different health programmes. |
| 2. Davis AM, Vinci LM, Okwuosa TM, Chase AR, Huang ES. Cardiovascular Health Disparities. Medical Care Research and Review. 2007;64(5_suppl):29S-100S. | Interventions were grouped by the vascular risk or condition they address (e.g. hypertension and hyperlipidaemia) and by the predominant target of the intervention: patient and family, provider and health care organizations, or multitarget interventions. | Condition-specific: 1) Hypertension- BP, dietary sodium, weight loss, BMI, cholesterol; 2) Hyperlipidaemia- weight loss, BMI, cholesterol, BP; 3) Tobacco consumption- quit rates, abstinence rates, readiness to quit; 4) Physical inactivity- step count, nutrition, exercise, health status, health behaviour, self-efficacy, emergency department visits, cardiovascular function and strength; 5) Congestive heart failure- daily weight, hospital admissions, quality of life. | Few studies specifically tested interventions for their effectiveness in reducing ethnic/racial disparities in cardiovascular prevention and care (no white subpopulation for comparison). Management and outreach strategies involving nurses have shown relatively consistent effectiveness in communities of colour. Pharmacist-led interventions for hypertension and hyperlipidaemia were relatively effective. Potassium supplementation may be of value in African communities. |
| 3. Engberg, M. E. (2004). "Improving intergroup relations in higher education: A critical examination of the influence of educational interventions on racial bias." Review of Educational Research 74(4): 473-524. | Multicultural course interventions (e.g. mandatory/ non-mandatory diversity courses; ethnic studies and women's studies); diversity workshop and training interventions; Peer-facilitated interventions (e.g. peer training, learning communities, collaborations, intergroup dialogue); Service interventions (e.g. community service, volunteer work). | Quantitative, qualitative and mixed methods measures of racial bias (prejudice, stereotypes, affective reactions, discrimination) | The majority of educational interventions are effective in reducing racial bias. As a whole, differential effects were found most often across different racial groups. In particular, White students were found to benefit more than students of colour, especially across diversity workshop and training interventions. Similarly, gender effects were found in a small number of studies, with women usually experiencing stronger effects or making greater change during the course. |
| 4. Escriba-Aguir, V., et al. (2016). "Effectiveness of patient-targeted interventions to promote cancer screening among ethnic minorities: A systematic review." Cancer Epidemiology 44: 22-39. | Quality improvement (QI) interventions to promote cancer screening among ethnic minorities delivered via the healthcare system and targeting the patients (not health professionals or health services). Interventions by cancer-type: 1) Colorectal cancer (6/17): 4/6 in community; 1 in primary care; 1 at home, all included educational component, peer education, free tests, reminders, small media, all culturally adapted; 2) Cervical (3/17): community or homes; education, counselling, small media, free test/navigation, reminders, all culturally adapted; 3) Breast (4/17) - range of interventions from guideline recommendations, all culturally adapted; 4) Cervical and breast (3/17); 5) Breast, colorectal, prostate, lung (1/17), culturally adapted small media materials by post. | Several variables related to screening: a) screening status (primary outcome: screening participation), b) knowledge awareness, self-efficacy, attitudes, intentions and perceptions, and c) health beliefs and barriers. | Results show that peer-based education on culturally targeted patient interventions may enhance effectiveness of interventions. Findings also suggest that the effectiveness of the interventions appears to be associated with the use of small media, one-on-one interactions, and small group education sessions for colorectal, breast and cervical cancer. In addition, patient reminder strategies seem to reinforce the effectiveness of interventions. Conclusions cannot be made about effect on racial disparities as there was no comparison with the White population. |
| 5. Glick, S., Clarke, A. R., Blanchard, A., et al. 2012. Interventions to improve minority health and reduce racial and ethnic disparities in care for cervical cancer: A systematic review. Journal of General Internal Medicine 2) S233. | Interventions to improve screening included patient educational materials and education programs, navigation (scheduling appointments, low-cost sources of care and transportation), low-cost screening, improved access to screening, reminders for healthcare providers, advertisements, office policies and procedures (such as protocols or tracking systems), telephone counselling or support, feedback for providers on screening rates, upgraded equipment. Interventions to improve diagnosis and treatment of premalignant disease included telephone counselling, a streamlined process for follow-up of abnormalities, intensive follow-up and/or vouchers for reduced cost care, a personalized letter, pamphlet, audiovisual presentation or transportation incentives, written educational materials, education programs, and messages in the media. | Rate of screening, ratio of in situ to invasive carcinomas. | There is moderate evidence for telephone support with navigation in increasing the rate of screening. There is a low strength of evidence for education delivered by lay health educators with navigation. There is insufficient evidence for all the other interventions and combinations of interventions. There is a low strength of evidence for the interventions to prevent and treat premalignant disease (telephone counselling and follow-up). |
| 6. T. L. Fisher, D. L. Burnet, E. S. Huang, M. H. Chin and K. A. Cagney. Cultural leverage: interventions using culture to narrow racial disparities in health care. Medical Care Research & Review 2007 Vol. 64 Issue 5 Suppl Pages 243S-82S. | The interventions clustered into three broad categories: (1) interventions that modified the health behaviours of individuals (14/38) e.g. community-crafted culturally-specific messages to change behaviour and health professionals or peers health professionals who were culturally specific to the targeted racial groups providing leadership in diet education, breast self-exam and asthma self-regulation; (2) interventions that increased access from communities to the existing health care environment e.g. screening programs incorporating patient navigators and lay educators who increased awareness and understanding of cancer screening; and (3) interventions that modified the health care system to better serve patients and their communities (10/38) nurses, counsellors, and community health workers to deliver culturally tailored health information | 6 of 38 studies did not report on health outcomes. The remaining 32 studies did report on health outcomes. 16 gathered outcome data from patient self-report, and 16 used body habitus, metabolic, and/or cardiovascular parameters. Other outcome measures included: improvements in asthma self-management, improvement in rates of cancer screening, decreased emergency department visits and hospitalizations in persons with diabetes, and developmental improvement in the children of the African American and Latina women. | Only a limited number of the interventions assessed health outcomes, and the demonstrated effect was not robust. Some common themes emerged: 1) nurses and other nonphysician health care providers implemented most of these interventions; 2) Those initiated by physicians were generally brief in duration and focused on training physicians in cultural tools or language acquisition; 3) Nurse-led studies often described in detail the extent to which race and ethnicity influence health care delivery. |
| 7. S. H. Hankerson and M. M. Weissman. Church-based health programs for mental disorders among african americans: A review. Psychiatric Services 2012 Vol. 63(3) Pages 243-249 | Church-based health promotion programs (support groups, focus groups and educational sessions) for substance-related disorders and anxiety and depressive symptoms. | Alcohol use, drug use, HIV/AIDS knowledge and attitudes, smoking cessation, stages of change, exposure to intervention, treatment attendance, treatment adherence, smoking quit rates, depressive symptoms (e.g. Beck Depression Inventory), problem recognition, decision to seek help, selection of helpers, satisfaction with support group, knowledge and understanding of mental illness and services. | Some promising results (reduced drug use, increased smoking cessation, improved depressive symptoms, improved knowledge and understanding of mental illness and services, improved treatment adherence). But evidence extremely limited due to the small number of studies, small numbers of participants, different types of data and lack of meta-analysis to assess the effectiveness of interventions. |
| 8. N. Hassen, A. Lofters, S. Michael, A. Mall, A. D. Pinto and J. Rackal. Implementing anti-racism interventions in healthcare settings: A scoping review. International Journal of Environmental Research and Public Health 2021 Vol. 18(6) Pages 1-15 | Individual level (cultural competency training, continuous ongoing training with anti-racism focus, critical reflection on knowledge, attitudes and beliefs); Interpersonal level (reflective questions on culturally safe healthcare, guidelines on how to address racist comments in psychotherapy); Community level (develop ongoing meaningful partnerships, actively engage racialised communities throughout the process, reorganise power by creating caucus groups for community organising), Organisational level (strategic leadership, commitment from leadership, policy compliance procedures, support for minority staff, a range of educational outputs, incorporate antiracism into quality improvement initiatives); Policy level (workforce policy to recruit, retain and promote minority staff, increase infrastructure, accountability, transparency and monitoring, shared leadership and decision making, mandate targets and actions, implement multilevel actions). | Experiences of workshops, change in attitudes, cross-cultural preparedness, interviews and observations. | Few studies included complete evaluation findings. Key considerations for anti-racism work in healthcare settings: 1) Define the problem and set clear goals and objectives; 2) Incorporate explicit and shared anti-racist language; 3) Establish leadership buy-in and commitment; 4) Invest dedicated funding and resources; 5) Bring in the right support and expertise (skilled facilitators, members of racialised groups). Establish ongoing, meaningful community and patient partnerships. Strategies for implementation and evaluation: 1) Use multi-level long term approach; 2) Embed racial equity policies and procedures (hiring, retention, and promotion); 3) Link mandatory anti-racism work to broader systems of power, hierarchy, and dominance; 4) Build in Stop and reflect mechanisms in a cyclical process. |
| 9. T. Jones, E. A. Luth, S. Y. Lin and A. A. Brody. Advance Care Planning, Palliative Care, and End-of-life Care Interventions for Racial and Ethnic Underrepresented Groups: A Systematic Review. *Journal of Pain and Symptom Management* 2021 Vol. 62(3) Pages e248-e260 | Educational interventions (n=13) to patients or caregivers to improve utilisation or access (language, knowledge attitude, beliefs, health literacy and culturally appropriate information delivery); Community Support (n=2) including peer or patient navigators to empower patients and offer support in the community (aiming to increase access and reduce mistrust); Clinical homebased palliative care to increase access and provide culturally sensitive care. | Knowledge, self-efficacy and skills related to EOL decision-making, intention to complete advanced care plan, experiences with advanced care planning, advance care plan documentation in the medical record, patient-surrogate congruence in EOL treatment preferences, patient and surrogate, comfort in decision-making, and psychosocial and spiritual well-being, quality of life, emergency department attendance, preferences for end of life care, length of hospice stay. | There was significant variation in outcomes making it difficult to compare effectiveness. Education interventions were most common but limitations need to be considered in individual level interventions implemented in diverse populations. The two studies involving peer navigation interventions were either a pilot trial or lacked a comparison group and did not show high effectiveness. |
| 10. S. Y. Lee-Tauler, J. Eun, D. Corbett and P. Y. Collins. A systematic review of interventions to improve initiation of mental health care among racial-ethnic minority groups. *Psychiatric Services* 2018 Vol. 69(6) Pages 628-647 | Interventions identified included: 1) collaborative care (N=10), 2) psychoeducation (N=7) which aimed to educate patients about psychotherapy entry and attendance, improve depression or psychosis literacy, reduce stigma associated with antidepressants, and increase access to help seeking, 3) case management (frequent and intensive contacts, small caseloads, and assertive outreach) (N=5), 4) colocation of mental health services comparing integrated care intervention where mental health services were located in primary care with enhanced referral model where psychiatric services were provided in a separate location (N=4), 5) screening and referral (N=2) which involved social workers and primary care providers trained to screen for depression, and 6) a change in Medicare medication reimbursement policy (the federal program that subsidizes the costs of prescription drugs and the drug insurance premium), that served as a natural experiment (N=1). | Outcome measures included initial access to mental health services (number of visits and consultations), pharmacological treatment initiation as a result of the study, willingness to seek psychological or pharmacological treatment, initiation of psychotherapy or pharmacologic treatment. | Seven studies provided evidence that screening and referral, colocation of primary care and mental health services, and collaborative care interventions improved mental health outcomes and reduced disparities in initiation of care. Six of the seven interventions used an integrated care model and resulted in increased uptake of psychotherapy or antidepressant use among members of racial-ethnic minority groups compared with white participants. |
| 11. M. Loutfy, W. Tharao, C. Logie, M. A. Aden, L. A. Chambers, W. Wu, et al. Systematic review of stigma reducing interventions for African/Black diasporic women. Journal of the International AIDS Society 2015 Vol. 18 Issue 1 | Only two studies designed their interventions specifically for African American women. The other 3 studies included African/Black diasporic women in their studies of interventions which were not culturally-adapted. Interventions included: 1) a cognitive intervention designed to foster the reframing of traumatic events and reduce negative thoughts, 2) a behavioural intervention designed to enhance HIV-knowledge, coping, social support and psychological skills building of youth recently diagnosed with HIV, 3) a maternal HIV self-care symptoms management intervention designed to improve HIV-related knowledge, reduce emotional distress and to promote self-care and care seeking strategies among low-income African American mothers with HIV, 4) participatory educational exercises designed to counter misinformation around HIV, raise awareness of HIV-related stigma and develop skills to addressing stigmatizing situations within one's daily life. | All studies reported stigma outcomes: perceived HIV stigma (i.e. awareness of social devaluation, social rejection, diminished social identity and limited social opportunity attributed to stigma), internalized stigma (i.e. holding negative views of oneself), well-being including depressive symptomology, health-related quality of life, health distress, number of infections and mood and affective state, disclosure self-efficacy, sexual discussion self-efficacy and coping, cognitive reorganization, HIV symptomology, adherence behaviours, side effects from antiretroviral drugs and HIV/AIDS knowledge. | 4/5 reported reductions in stigma post-intervention. Only study with a gender and ethno-racially diverse sample demonstrated mixed results in regards to stigma reduction (stigma and negative self-perception increased for women but decreased for men). Findings on physical and mental well-being post-intervention were mixed (depressive symptoms, self-efficacy, and social support). |
| 12. Marshall, S, Taki, S, Laird, Y, Love, P, Wen, LM, Rissel, C. Cultural adaptations of obesity-related behavioral prevention interventions in early childhood: A systematic review. *Obesity Reviews*. 2022; 23(4):e13402. doi:10.1111/obr.13402 | Most cultural adaptations involved modifying language and translations, altering activities to improve suitability, addressing cultural values in the intervention content, and involving culturally matched intervention facilitators. | Health and behavioural outcomes e.g. body mass index [BMI], neck circumference, and diastolic blood pressure for parent/caregivers, television viewing time. | Results were mixed and only one intervention (Fit 5 Kids which used songs and poetry familiar to Latino families) showed evidence of significant change in behavioural outcomes (reduced television viewing time). The interventions were varied and the use of cultural adaptation guidance and involvement of target communities were inconsistent. More rigorous and consistent use of theory and reporting standards is needed. |
| 13. M. L. McPheeters, S. Kripalani, N. B. Peterson, R. T. Idowu, R. N. Jerome, S. A. Potter, et al. Closing the quality gap: revisiting the state of the science (vol. 3: quality improvement interventions to address health disparities). Evidence report/technology assessment 2012 Issue 208 .3 Pages 1-475 | Most interventions (n=14) involved patient education. Ten studies incorporated self-management for example, teaching people with diabetes to check their blood sugar regularly. Eight studies included health care provider education, either focussing on the clinical issue or raising awareness about disparities affecting the target population. Other intervention categories were: audit and feedback, facilitated relay of clinical data to providers, patient and provider reminder systems and organizational change. The review focused on the following clinical conditions: breast cancer, colorectal cancer, diabetes, heart failure, hypertension, coronary artery disease, asthma, major depressive disorder, cystic fibrosis, pneumonia, pregnancy, and end-stage renal disease. | Outcome measures included health outcomes (e.g., morbidity and mortality, blood pressure and HbA1c); process measures (e.g., proportion of patients treated according to clinical guidelines); changes in disparity; and harms (i.e., negative impact of the intervention on the individual patients or the health care system). | Most studies (n=8) were unable to show a reduction in disparity. Most studies have focused on racial or ethnic disparities, with some targeted interventions demonstrating greater effect in racial minorities—specifically, supporting individuals in tracking their blood pressure at home to reduce blood pressure and collaborative care to improve depression care. One disease management and patient education program was associated with a reduction in disparity between Black and White patients in HbA1c testing when it was targeted in a geographic area with very high rates of diabetes. |
| 14. E. L. Paluck, R. Porat, C. S. Clark and D. P. Green. Prejudice Reduction: Progress and Challenges. Annual review of psychology 2021 Vol. 72 Pages 533-560 | Extended and imaginary contact, cognitive and emotional training, social categorization, peer influence and dialogue, value consistency and self-worth, interpersonal contact, multicultural antibias moral education, diversity training, cooperative learning, intercultural training and conflict resolution. | Change in attitudes or beliefs, behaviour, emotion, empathy and perceived norms. | The review provides evidence of moderate effect, but effect sizes were limited in size, scope, or duration. Some studies found effects diminished over time or were localized to some types of participants. Several studies found effects only on some types of outcomes but not others. For example, prejudice reduction interventions were more effective at changing behaviours than attitudes. The studies demonstrated publication bias. Publication bias occurs when the direction or strength of a study's outcome influences whether it is published or not. The result is that effects were less when restricted to studies with larger ($n > 78$) samples (the size of the intervention effect drops as study size increases). In other words, if the current studies had been conducted on a much larger scale, the analysis would have shown no reduction in prejudice. |
| 15. Peek ME, Cargill A, Huang ES. Diabetes Health Disparities. Medical Care Research and Review. 2007;64(5_suppl):101S-156S. doi:10.1177/1077558707305409 | Interventions were categorized as 1) targeting the patient; 2) targeting the provider; 3) targeting the healthcare system; 4) multi-target interventions. Patient interventions involved culturally-tailored self-management education, nurse and dietician support, and group sessions. Provider interventions included provider education, practice guidelines, audit and feedback, decision-support tools and computerised reminders about medication titration. Healthcare provider interventions included culturally tailored case management, community clinics, pharmacist-led support, bilingual staff and treatment algorithms. Multitarget interventions included collaborative Quality Improvement (QI) programs, communication training and patient empowerment, outreach programs and patient incentives. | Diabetes processes of care (i.e., measurement of HbA1c, blood pressure, and cholesterol) and intermediate diabetes health outcomes (i.e., control of HbA1c, blood pressure, and cholesterol) | On average, the interventions improved quality of care, health outcomes (such as diabetes control and reduced diabetes complications), and possibly reduced health disparities in quality of care. There was no ideal setting and interventions targeting patients, providers, and health care organizations were all able to bring about improvements. Successful patient-targeted interventions used interpersonal (non computer-based) skills and social networks such as family members, peer support groups, and one-on-one education. Culturally tailored interventions were much more effective than generalized diabetes self-management training interventions. Successful provider interventions involved in-person feedback rather than computerized decision-support in order to effect sustained behavioural change and health improvements. The majority of successful health system interventions focused on innovative use of staff including nurse case management, community health workers, nonphysician clinicians, and staff-led prescription assistance programs. |
| 16. K. Schill and S. Caxaj. Cultural safety strategies for rural Indigenous palliative care: A scoping review. BMC Palliative Care 2019 Vol. 18(1) (no pagination) | Symbolic or small gestures; anticipating barriers to care; defer to client, family and community; shared decision-making; active patient and family involvement; respectful, clear, and culturally appropriate communication; community ownership of services; empower cultural identity, knowledge, and traditions; and, policy. | This is a descriptive review in the context of indigenous health. The review aims to identify pathways and models of care. | Culturally competent practices improve services but do not improve disparities. Strategies include: symbolic or small gestures; anticipating barriers to access; deferring to the client, family, and community members; and, collective decision making and family involvement. Culturally safe approaches contribute to organizational change and improving disparities. Strategies include: involvement of patient and family in service planning; reflection about individual and systemic racism; community ownership of services and; recognizing distinct Worldviews that shape care. |
| 17. M. Truong, Y. Paradies and N. Priest. Interventions to improve cultural competency in healthcare: a systematic review of reviews. BMC health services research 2014 Vol. 14 Pages 99 | Interventions to improve cultural competency were: training/workshops/programs for health practitioners (e.g. doctors, nurses and community health workers), culturally specific/tailored education or programs for patient/clients, interpreter services, peer education, patient navigators and exchange programs. | There were three main categories of study outcomes: patient-related outcomes, provider-related outcomes, and health service access and utilization outcomes. Provider outcomes focused on knowledge, attitudes and skills related to cultural competency. Patient outcomes included physiological measures such as blood glucose, weight and blood pressure as well as patient satisfaction and trust, knowledge of cancer screening and knowledge of health conditions. Behavioural outcomes such as dietary and exercise behaviours were also measured. Outcomes related to health service access and utilization included use of bilingual community health workers, interpreters, and patient navigators. Cost-effectiveness of interventions was considered in three reviews. | The majority of reviews found moderate evidence of improvement in provider outcomes and health care access and utilization outcomes but weaker evidence for improvements in patient/client outcomes. A lack of methodological rigor was common and many of the studies relied on self-report, which is subject to a range of biases. There was a lack of evidence of objective intervention effectiveness. |
| 18.<br>Waller. Broken fixes: A systematic analysis of the effectiveness of modern and postmodern interventions utilized to decrease IPV perpetration among Black males remanded to treatment. *Aggression and Violent Behavior* 2016 Vol. 27 Pages 42-49. Accession Number: 608556985 DOI: http://dx.doi.org/10.1016/j.avb.2016.02.003 | CBT, psychoeducation, Duluth model or gender based psychoeducation, goal-setting. Goal-setting interventions are strengths-based and developed to empower marginalized and disenfranchised populations who may be less educated, underemployed and stigmatized. It allows a shift in power to the client, allowing him to develop treatment goals and collaborate with the therapist. | Programme attrition/adherence and recidivism | Outcomes for Black males were worse (higher attrition rates). The interventions replicate the discrimination and disempowerment in society and therefore caused greater harm for Black male offenders who suffer from the oppression of racism and a compromised sense of agency. The most effective treatment for Black males are those that incorporate cultural nuances related to power, marginalization and differential educational levels within a cultural-historical context. In this respect, the goal-setting interventions show the most promise. in terms of evaluation, program developers and researchers need to collaborate on a standardized definition for program dropout to improve evaluation, less reliance on perpetrator self-report is needed. |

### Critical appraisal

Using the AMSTAR2 criteria, one study (9%) was assessed as high quality, six studies (36%) were low quality and 11 (61%) were critically low quality (see Appendix 2-3). Methodological strengths across the included reviews were a comprehensive literature strategy (n=16, 94%), study selection completed in duplicate (n=12, 71%), data extraction completed in duplicate (n=13, 76%), adequate description of included studies (n= 9, 53%) and conflicts of interests of authors declared (n=14, 82%). Methodological weaknesses across the studies were protocol registration (n= 4, 24%), risk of bias from individual studies included in the review (n= 3, 18%), meta-analytical methods (n= 1, 9%), consideration of risk of bias in interpretation of results (n= 1, 9%) and assessment of publication bias (n= 1, 9%). The AMSTAR2 tool is based on the AMSTAR tool which was designed for randomised controlled trials (RCTs) ^37^. The revised AMSTAR2 enables appraisal of both randomised and non-randomised studies of healthcare interventions but retains 10 of the original domains, including assessment of risk of bias from unconcealed allocation and lack of blinding, sources of funding of studies, and conduct of meta-analysis ^37^. This makes it less well suited to the studies included in this review, including studies where the comparator group is not clearly described and narrative syntheses due to heterogeneity of interventions.

### Results of individual sources of evidence

A review of the characteristics of the interventions presented in the included studies is described in Appendix 4, and a summary of the interventions with some evidence of effectiveness is presented in **Figure 3**.

**Figure 3.**
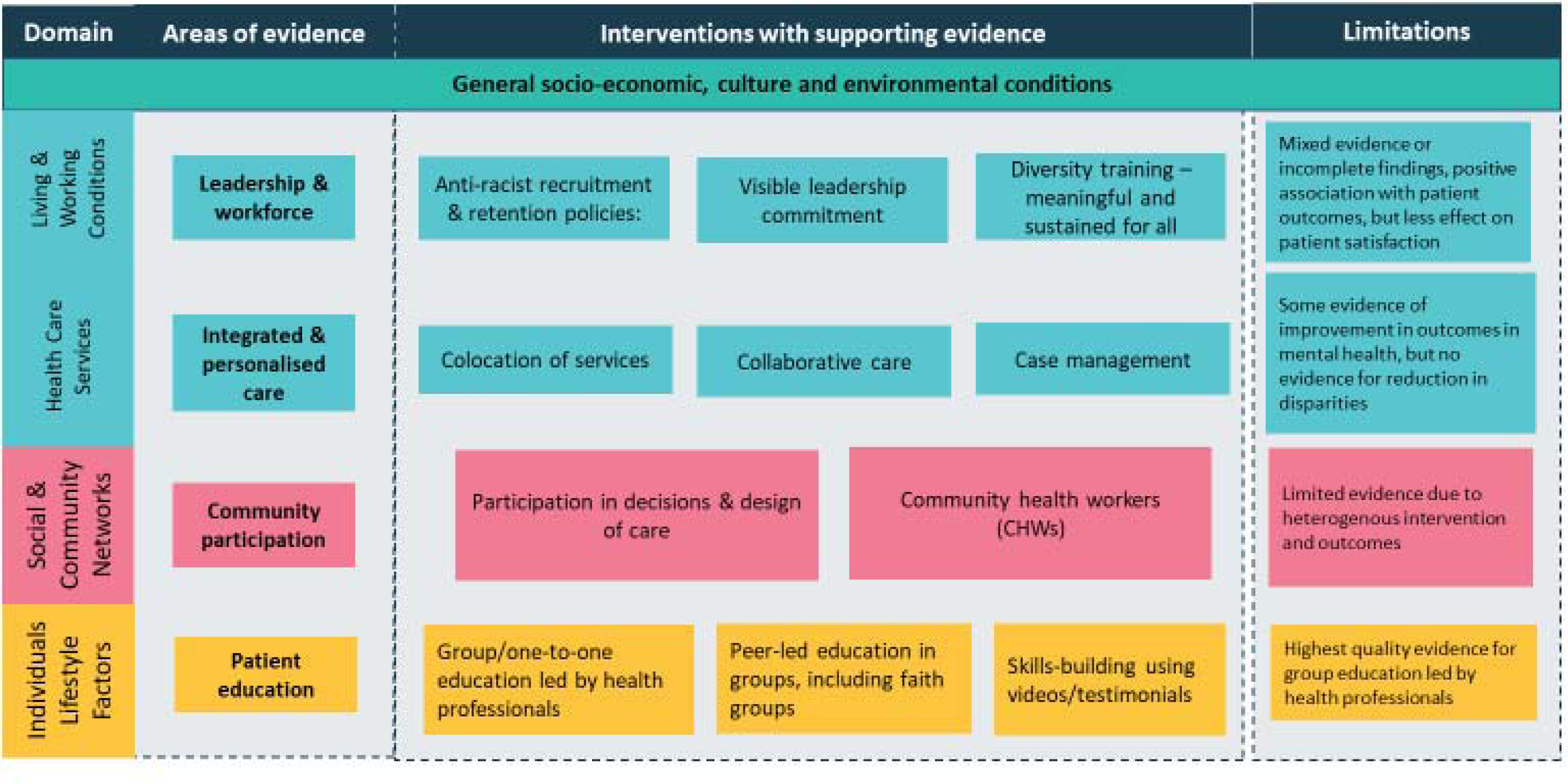
Summary of interventions with some evidence of effectiveness, and limitations of evidence, grouped by domains drawn from the socio-ecological model.

## Effectiveness of interventions

### Individual and community level interventions

#### Patient education and access

The evidence for interventions targeting education and access for ethnic minorities is mixed. A review of diabetes interventions found that on average, these educational interventions improved quality of care, health outcomes (such as diabetes control and reduced diabetes complications), and possibly reduced health disparities in quality of care ^47^. Effective patient-targeted interventions used interpersonal (non-computer-based) skills and social networks such as family members, peer support groups, and one-on-one education. **Culturally tailored interventions were more effective** than generalized diabetes self-management training interventions ^47^. The review of coalition-driven interventions found that **group-based health education led by professional staff** produced fairly consistent positive effects on health and behavioural outcomes ^53^. There were mixed results from group-based health education led by peers on behavioural outcomes, but church-based peer interventions in two studies resulted in improved BMI ^53^.

Most other studies of patient education interventions were unable to show evidence of a reduction in ethnic disparities. Limitations included health outcomes not being assessed, no white subpopulation for comparison, the demonstrated effect sizes not being robust, or lack of general conclusions about which interventions work for whom due to a wide range of interventions targeting different health programmes ^48 50 53 55^.

The evidence for church-based mental health programs is promising. Intervention components include emphasizing black culture and spirituality, using Churches as a setting, involving Church mentors, and including prayer. Outcomes include reduced drug use, increased smoking cessation, improved depressive symptoms, improved knowledge and understanding of mental illness and services, improved treatment adherence ^41^. However, the evidence is extremely limited due to the small number of studies, small numbers of participants, different types of data and lack of meta-analysis to assess the effectiveness of interventions ^41^.

Findings from the review of stigma-reducing interventions (including patient education to increase HIV knowledge and address misinformation) for African and Black diasporic women were mixed ^45^. Four out of five of the included studies demonstrated a positive effect on reducing HIV-related stigma in women living with HIV ^45^. However only two of these studies exclusively sampled African and Black diasporic women and developed interventions culturally appropriate for this population ^45^.

Reviews of cancer interventions found low strength or insufficient evidence for patient education ^39 40^. A review of cancer-screening interventions found that peer-based education may enhance effectiveness of interventions, and this appears to be associated with the use of small media, one-on-one interactions, and small group education sessions ^39^. Patient education interventions had better strength of evidence than access interventions including free tests, support with navigating appointments and appointment reminders ^39^. However limitations of studies of cancer interventions included lack of comparison with the White population, reliance on self-reporting, and lack of statistically significant differences^39 40^.

Reviews of palliative care interventions found that there was significant variation in types of outcomes used, making it difficult to compare effectiveness ^43^.

##### Cultural adaptation

The evidence for **culturally adapted education interventions** for palliative care is promising ^43^. Most interventions had significant associations with improved patient engagement, change in attitudes and knowledge of advanced care planning, and congruence in goals of care. One high quality randomised controlled trial (RCT) found that a multi-lingual online interactive skill-building program designed especially for diverse patients and carers using video stories, narratives, and testimonials to model how to engage in advanced care planning significantly increased documentation of advanced directives and engagement with advanced care planning when the intervention was compared to non-culturally adapted easy-to-read advanced directives ^43^.

*Though results for patient and community-based education to improve health outcomes for ethnic minority groups, and/or reduce ethnic health disparities are mixed; the most promising interventions reviewed include group-based health education led by professional staff and providing culturally tailored or adapted interventions. These interventions were supported by the highest quality evidence and were most consistent amongst studies reviewed*.

###### Community partnership-building

Reviews which included interventions targeting community partnership-building found that these interventions **contributed to organizational change and improving disparities** ^44^. Strategies include involvement of patients and families in service planning, **reflection about individual and systemic racism** community ownership of services and recognizing distinct world views that shape care ^44^.

##### Lay Community Health Workers

Lay Community Health Workers (CHW)s compared favourably with broad-scale community and health system interventions ^53^. Results from the review of coalition-driven interventions suggested beneficial changes in health behaviour and health status measures when CHWs provided support, but results were *not consistent*across studies and were quality appraised as ‘low-certainty evidence’ ^53^. Evidence for CHWs in cardiovascular disease is promising. A study of enhanced tracking and follow-up of low-income African Americans with hypertension by CHWs found that clinic attendance improved from 47% to 65% compared to usual care ^55^. Another study demonstrated a 50% reduction in emergency department attendances after employing Community Health Workers to work with a group of diabetic hypertensive patients ^55^. However, Community Health Worker interventions were found to be *heterogenous* in terms of approach and outcome measurement. There was little description of the training or characteristics of the CHWs or how this impacted their success. Many studies focused on improved understanding of disease or satisfaction with care, and trends toward improving health behaviours rather than change in health behaviour itself ^48^.

*Community partnership building, cultural adaptions and community health workers are different forms of participation in the decisions, design and delivery of health services. Community participation is considered a driver of health equity.* ^56^ *Taken together, these studies suggest that participation in all aspects of care pathway development that empowers ethnic minority groups may provide an effective approach to reducing ethnic health disparities*.

#### Healthcare organisation level interventions

Healthcare organisation level interventions can be grouped into three categories: organisation of care, clinician interactions with patients, and workforce and leadership.

##### Organisation of care

Interventions targeting organisation of care (collaborative care, case management and colocation of services) were described in two reviews ^42 50^. In a review of interventions to improve initiation of mental health care, seven out of twenty-nine studies provided evidence **that colocation** of primary care and mental health services and collaborative care interventions not only improved mental health outcomes but also contributed to disparities reduction in initiation of care ^42^. Findings included increased uptake of psychotherapy or antidepressant use among members of ethnic minority groups compared with white participants ^42^. A second review found that collaborative care resulted in greater effects in ethnic minority patients with depression, including depression scores, severity, and functioning ^50^. Collaborative care was more effective in less-educated individuals than in those with more education and in women than in men for some care outcomes in patients with depression ^50^. However none of the studies demonstrated a specific reduction in disparity caused by the intervention, partly because few disparities were measurable at baseline ^50^.

*These studies show that different types of pathway integration, and providing a more person-centred approach with fewer steps for patients to navigate can contribute to reducing disparities*.

##### Clinician interactions with patients

A review of diabetes health disparities found a number of studies targeting clinician behaviour, the majority of which involved the application of generic diabetes quality improvement initiatives to ethnic minority groups ^47^. The interventions typically included practice guidelines, continuing medical education, computerized decision-support reminders, in-person feedback and problem-based learning. The interventions resulted in improved processes of care (HbA1c monitoring, foot care, exercise counselling etc) and improved diabetes control. None of the interventions included culturally tailored components ^47^. However improved care and control is particularly relevant to ethnic minorities as evidence suggests they are less likely to have access to care and more likely to have worse control of their diabetes ^47^. This suggests **that targeting clinicians for quality improvement in service delivery in areas with higher proportions of ethnic minority populations may be an effective strategy** for improving diabetes outcomes in these groups ^47^. Such targeted approach to a general condition or service provision is in line with a proportionate universalism approach to address health inequalities. ^57^

##### Workforce and leadership

Evidence for workforce and leadership interventions is lacking due to methodological issues in individual studies. One review describing interventions targeting workforce and leadership (diversity training, leadership quality improvement initiatives, and recruitment and retention policies) found that few studies had complete evaluation findings ^49^. A second review found mixed evidence for staff diversity and cultural competency training, with a positive relationship between cultural competency training and improved patient outcomes, but less effect on patient satisfaction with care ^52^.

*The reviews found that overall there is an overemphasis on individual-centred education and individual behaviour change rather than organizational change, and recommend that focus and resources shift to policies and practices that seek to dismantle institutional and systemic racism through a multi-level approach, where cultural competency training is only one component and not a standalone intervention.*^52^ The studies also show that better and more consistent data collection and research methods are required to improve evidence on workforce and leadership training.

Tools and resources for addressing organizational racism have been identified from the education and non-profit sectors and we describe evidence from outside the healthcare sector below.

##### Interventions outside the healthcare setting

###### Education

A review by Engberg et al found that most education interventions in the included studies were effective at **reducing racial bias** ^34^. The evidence was stronger for ethnic and women’s’ studies courses (long-term interventions) than diversity workshops (short-term interventions), and white students were found to benefit more than students of colour ^34^. Several limitations of the included studies were highlighted including lack of scales to measure racial bias accurately, reliance on quasi experimental study designs that are vulnerable to selection bias and reliance on convenience or purposive sampling, which limit generalizability to other populations ^34^.

A review of prejudice reduction interventions in higher education (including antibias, diversity and intercultural training) found evidence of moderate effect, but effect sizes were limited in size, scope, or duration ^54^. Some studies found effects diminished over time or were localized to some types of participants. Several studies found effects only on some types of outcomes but not others. For example, prejudice reduction interventions were more effective at changing behaviours than attitudes. The studies demonstrated publication bias, and if the studies had been conducted on a larger scale, the analysis would have shown no reduction in prejudice ^54^.

###### Criminal justice

A review of interventions to decrease intimate partner violence perpetration among Black males remanded to treatment found that outcomes for Black males were worse than for their white counterparts ^36^ due to the interventions mirroring societal discrimination ^36^. The most effective treatments for Black males are those that **incorporate cultural nuances related to power**, marginalization and differential educational levels, with co-developed goal-setting interventions showing the most promise ^36^.

## Discussion

We found two levels of interventions based on the socioecological model (Figure 1), one operating at the institutional level and one located at the community and individual level. We found that many of the interventions in service delivery target individuals and involve education, and though the results for educational interventions were mixed, group-based health education led by professional staff and providing culturally tailored interventions were supported by the highest quality and most consistent evidence to reduce ethnic inequalities. Culturally tailoring interventions, together with collaborative community partnerships (community health workers and service user participation in developing and delivering health services), provides agency to disadvantaged groups and allows them to contribute to services that meet their needs. Empowerment and inclusion were also evident in the systematic review to reduce recidivism, without which there is potential to increase inequalities in all aspects of planning and delivery of care ^58^.

Integrated care with models of collaboration between different disciplines, co-location of services and case management to provide a more patient centred approach, was also identified as effective in treatment for minority ethnic groups, but did not reduce inequalities due to lack of baseline measurement. Complex systems can be difficult to navigate, particular for those with fewer resources or language constraints. Simplifying access through integrated services can overcome some of the barriers.

For organisational interventions, the reviews found that overall, there is an overemphasis on patient education and individual behaviour change rather than organizational change and recommend shifting focus and resources to policies and practices which seek to dismantle institutional and systemic racism through a multi-level approach, where cultural competency training is only one component and not a standalone intervention. Other components would include ensuring a leadership commitment, a range of workforce interventions to address unfair recruitment, retention and promotion practices, and anti-racist quality improvement initiatives.

### Comparison to previous literature

There have been an several publications in recent years on tackling racism in health and care, but not all were systematic reviews, and some did not explicitly aim to address ethnic health inequalities. Hassan et al,^59^ one of the included studies, conducted a scoping review on anti-racist interventions in healthcare settings, and included 12 empirical studies and 25 conceptual papers, in contrast to this study, which only selected systematic reviews of empirical studies with interventions and took account of potential impact on ethnic disparities. Though we drew similar conclusions, we provide more detail on specific interventions that could be employed to address ethnic disparities through an anti-racist approach.

A recent Lancet series on racism, xenophobia, discrimination, and health applied an anti-racist frame on global health and health inequalities. ^7 20^ The authors recommend six strategic principles to address related health harms, which includes a decolonisation perspective, addressing both reparative and transformative justice, increasing diversity and inclusion to improve social cohesion and resilience, including considerations of intersectionality, taking an anti-racist approach at all socioecological levels, and also supporting human-rights based approaches. There was less emphasis on community participation compared to our findings. We took a narrower approach to distil key learning that organisations could action, but would take the historical, intersectional and rights considerations to contextualise how actions and interventions could be taken forward. Additionally, increasing diversity and inclusion in the workforce, and acting in all levels is also highlighted in our review and framework.

Overall, this study strengthens and integrates a range of previous studies to provide an evidence base for organisations to take an anti-racist approach to address ethnic health disparities.

### Strengths and Limitations

This study employed a rigorous review process with a comprehensive search strategy across multiple databases, and dual independent review. Our categorisation and synthesis of results used the socioecological model, which worked well to understand the different levels in which the interventions were acting. There were also a number of limitations to our study. Firstly, although we selected systematic reviews, many were considered low or critically low in quality due to issues in reporting, such as not reporting the reviewed studies’ funding sources, or listing excluded studies, and methodological limitations, including small sample sizes, and lack of direct comparators. We note that appraisal tools to assess the quality of reviews are based on biomedical standards, derived from a reductionist frame to infer causality in scientific studies. Using this lens can downgrade the value we place on studies of health that draws on social factors, such as race equity. Designing instruments that takes into account both social science and biomedical science perspectives is an important development in this area.

We limited reports to peer-reviewed literature in English, to facilitate access, but this may have reduced the scope of our findings. We may have also missed important findings from other sources. Therefore relevant literature that were identified during the selection process, but did not meet the inclusion criteria were also reviewed and considered alongside the selected studies to provide context and additional insights which may have mitigated this risk. Publication bias may have also resulted in positive results and we cannot rule out missing studies. Heterogeneity of intervention approaches, study designs, and reporting presented in the included articles made comparing results difficult. For the individual interventions, there was limited reporting of cultural adaptation, implementation and also lack of comparison with White ethnic groups, which limited our understanding of the impact on ethnic health inequalities.

### Implications for Policy and practice

Health and care systems are keen to act on ethnic health inequalities, and have already implemented the NHS workforce race equality strategy and metrics, with the London health and care system going even further in their support of the London workforce. ^60^ Based on this review, we recommend five areas of action for health and care organisations:

1. A leadership commitment to infuse policies and practices with an anti-racist lens, including embedded and sustained training for the whole organisation to ensure cultural competence for the population served.
2. Supporting ethnic minority workforce and addressing racism in recruitment and retention policies.
3. Providing health programmes based on integrated patient centred care with an anti-racism focus, for example, collaborative care and colocation of services.
4. Many health and care organisation are anchor institutions, which can play a significant role in the social, economic and environmental conditions of communities within which they are situated. Commitment to anchor principles, particularly from areas with high proportions of ethnic minority groups can support local ethnic minority communities.
5. Community participation in the decisions, design, delivery and evaluation of services. Building trust, and capacity for communities to participate can support efforts to reduce health inequalities.

## Conclusions

This scoping review represents, to our knowledge, the first examination of interventions to address ethnic health disparities with an anti-racist approach. Despite the limitations in the quality of evidence from a biomedical quality assessment tool, there are strong themes evident in the studies reviewed and from other sources of published literature identifying effective approaches to addressing ethnic health inequalities. Given the urgent nature of this long-standing issue, we have made five recommendations for policy and practice for health and care organisations to start their work in embedding an anti-racist approach to tackling ethnic health inequalities. Our recommendations are not a complete list of activities, but a strategic framework from which to start building programmes in collaboration with communities. Individual-based approaches that target discrimination though important, will not be sufficient to reduce the impact of racism is all its forms, and progress towards racial health equity. The systemic and structural nature of racism will require organisations and systems to change, embedding an anti-racist lens in all policies and programmes of work.

The interventions outlined can help organisations to make a start on tackling ethnic health disparities – however, this is about “what” we do, not “how” we do it. We also recognise the need to take decolonisation, social justice, intersectionality and trauma-informed approaches in anti-racism, and these perspectives infuses “how” the work is done.

There was a lack of clear evidence on impact on ethnic health inequalities in the majority of the studies, due to lack of baseline or comparison with White ethnic groups. Planned evaluation and better data collection is an important consideration for next steps, including better co-ordination between health care providers to allow more standardised ways of reporting outcomes and processes to understand impact on communities.

## Data Availability

All data produced in the present study are available upon reasonable request to the authors

## Contributors

JY and KF conceived and designed the study. JY and SC designed the database searches and carried out the searches. JY, SC and LDT screened titles and abstracts for inclusion. JY, SC and SP conducted data extraction. JY and SP analysed the data and drafted the manuscript. All authors contributed to interpretation of the results, provided critical review and approved the final manuscript. SP is the guarantor and accepts full responsibility for the work and/or the conduct of the study, had full access to all the data in the study and takes responsibility for the integrity of the data and the accuracy of the data analyses.

The corresponding author attests that all listed authors meet authorship criteria and that no others meeting the criteria have been omitted.

## Ethics approval

Not required

## Acknowledgements

We would like to thank Jennifer Ford UK Health Security Agency Librarian, and the London Health Equity Group members for their feedback in the early stages of the work.

## REFERENCES

1. Office of Health Improvement and Disparities DoH. Official Statistics Health Inequalities Dashboard: statistical commentary, June 2022 2022 [Available from: https://www.gov.uk/government/statistics/health-inequalities-dashboard-june-2022-data-update/health-inequalities-dashboard-statistical-commentary-june-2022 accessed 30/1/2023 2023.

2. Taylor R, Rieger A. Medicine as social science: Rudolf Virchow on the typhus epidemic in Upper Silesia. International journal of health services: planning, administration, evaluation 1985;15(4):547–59.

3. Department of Health and Social Security. Report of the Working Group on Inequalities of Health. London, 1980.

4. Department of Health. Independent Inquiry into Inequalities in Health Report. London, 1998.

5. World Health Organisation. Commission on Social Determinants of Health - final report. Geneva, 2008.

6. Marmot M AJ, Boyce T, Goldblatt P, Morrison J. Health Equity in England: The Marmot Review 10 Years On. 2020

7. Devakumar D, Selvarajah S, Abubakar I, et al. Racism, xenophobia, discrimination, and the determination of health. Lancet 2022;400(10368):2097–108. doi: 10.1016/S0140-6736(22)01972-9

8. Mathur R, Rentsch CT, Morton CE, et al. Ethnic differences in SARS-CoV-2 infection and COVID-19-related hospitalisation, intensive care unit admission, and death in 17 million adults in England: an observational cohort study using the OpenSAFELY platform. Lancet 2021;397(10286):1711–24. doi: 10.1016/S0140-6736(21)00634-6 [published Online First: 20210430]

9. Nafilyan V, Islam N, Mathur R, et al. Ethnic differences in COVID-19 mortality during the first two waves of the Coronavirus Pandemic: a nationwide cohort study of 29 million adults in England. Eur J Epidemiol 2021;36(6):605–17. doi: 10.1007/s10654-021-00765-1 [published Online First: 20210616]

10. Public Health England. Disparities in the risk and outcomes of COVID-19, 2020.

11. Williamson EJ, McDonald HI, Bhaskaran K, et al. Risks of covid-19 hospital admission and death for people with learning disability: population based cohort study using the OpenSAFELY platform. BMJ 2021;374:n1592. doi: 10.1136/bmj.n1592 [published Online First: 20210714]

12. Office for National Statistics (ONS). Updating ethnic contrasts in deaths involving the coronavirus (COVID-19), England: 8 December 2020 to 1 December 2021. Office for National Statistics, 2022.

13. Office of Health Improvement and Disparities (OHID) DoHaSC. COVID-19 Health Inequalities Monitoring for England (CHIME) tool 2023 [Available from: https://analytics.phe.gov.uk/apps/chime/ accessed 30/1/2023 2023.

14. Hoernke K, Djellouli N, Andrews L, et al. Frontline healthcare workers’ experiences with personal protective equipment during the COVID-19 pandemic in the UK: a rapid qualitative appraisal. BMJ Open 2021;11(1):e046199. doi: 10.1136/bmjopen-2020-046199 [published Online First: 20210120]

15. Paradies Y, Ben J, Denson N, et al. Racism as a Determinant of Health: A Systematic Review and Meta-Analysis. PLoS One 2015;10(9):e0138511. doi: 10.1371/journal.pone.0138511 [published Online First: 20150923]

16. MBRRACE-UK. Saving Lives Improving Mothers’ Care - Lessons learned to inform maternity care from the UK and Ireland Confidential Enquiries into Maternal Deaths and Morbidity 2018-20. 2022

17. Matthews RJ, Draper ES, Manktelow BN, et al. Understanding ethnic inequalities in stillbirth rates: a UK population-based cohort study. BMJ Open 2022;12(2):e057412. doi: 10.1136/bmjopen-2021-057412 [published Online First: 20220309]

18. Kapadia D, Zhang J, Salway S, et al. Ethnic Inequalities in Healthcare: A Rapid Evidence Review: NHS Race and Health Observatory, 2022.

19. Selvarajah S, Corona Maioli S, Deivanayagam TA, et al. Racism, xenophobia, and discrimination: mapping pathways to health outcomes. Lancet 2022;400(10368):2109–24. doi: 10.1016/S0140-6736(22)02484-9

20. Abubakar I, Gram L, Lasoye S, et al. Confronting the consequences of racism, xenophobia, and discrimination on health and health-care systems. Lancet 2022;400(10368):2137–46. doi: 10.1016/S0140-6736(22)01989-4

21. Munn Z, Peters MDJ, Stern C, et al. Systematic review or scoping review? Guidance for authors when choosing between a systematic or scoping review approach. BMC Med Res Methodol 2018;18(1):143. doi: 10.1186/s12874-018-0611-x [published Online First: 20181119]

22. Kendi IX. How to be an anti-racist.: One World 2019.

23. Powell JA. Structural Racism: Building upon the Insights of John Calmore. North Carolina Law Review 2008;86(3):791–815.

24. Bonilla-Silva E. Rethinking Racism: Toward a Structural Interpretation. American Sociological Review 1997;62(3):465–80.

25. Harrell SP. A multidimensional conceptualization of racism-related stress: implications for the well-being of people of color. Am J Orthopsychiatry 2000;70(1):42–57. doi: 10.1037/h0087722

26. Hicken MT, Kravitz-Wirtz N, Durkee M, et al. Racial inequalities in health: Framing future research. Soc Sci Med 2018;199:11–18. doi: 10.1016/j.socscimed.2017.12.027 [published Online First: 20180102]

27. Williams DR, Lawrence JA, Davis BA. Racism and Health: Evidence and Needed Research. Annu Rev Public Health 2019;40:105–25. doi: 10.1146/annurev-publhealth-040218-043750 [published Online First: 20190202]

28. Gee GC, Ford CL. STRUCTURAL RACISM AND HEALTH INEQUITIES: Old Issues, New Directions. Du Bois Rev 2011;8(1):115–32. doi: 10.1017/S1742058X11000130

29. Calliste AM, Dei GJS. Power, Knowledge and Anti-Racism Education: A Critical Reader2000.

30. Farr M. Power dynamics and collaborative mechanisms in co-production and co-design processes. Critical Social Policy 2018;38(4):623–44.

31. Yip JL, DeSouza Thomas L. Beyond the conversation about race Better Health For All, 2021.

32. Dahlgren G, Whitehead M. The Dahlgren-Whitehead model of health determinants: 30 years on and still chasing rainbows. Public Health 2021;199:20–24. doi: 10.1016/j.puhe.2021.08.009 [published Online First: 20210914]

33. Tricco AC, Lillie E, Zarin W, et al. PRISMA Extension for Scoping Reviews (PRISMA-ScR): Checklist and Explanation. Ann Intern Med 2018;169(7):467–73. doi: 10.7326/M18-0850 [published Online First: 20180904]

34. Engberg ME. Improving intergroup relations in higher education: A critical examination of the influence of educational interventions on racial bias. Review of Educational Research 2004;74(4):473–524. doi: 10.3102/00346543074004473

35. Paluck EL, Porat R, Clark CS, et al. Prejudice Reduction: Progress and Challenges. Annual review of psychology 2021;72:533–60. doi: http://dx.doi.org/10.1146/annurev-psych-071620-030619

36. Waller B. Broken fixes: A systematic analysis of the effectiveness of modern and postmodern interventions utilized to decrease IPV perpetration among Black males remanded to treatment. Aggression and Violent Behavior 2016;27:42–49. doi: http://dx.doi.org/10.1016/j.avb.2016.02.003

37. Shea BJ, Reeves BC, Wells G, et al. AMSTAR 2: a critical appraisal tool for systematic reviews that include randomised or non-randomised studies of healthcare interventions, or both. BMJ 2017;358:j4008. doi: 10.1136/bmj.j4008 [published Online First: 20170921]

38. Davis AM, Vinci LM, Okwuosa TM, et al. ACCEPT Cardiovascular health disparities: A systematic review of health care interventions. Medical Care Research and Review2007;64(5 SUPPL.):29S–100S. doi: http://dx.doi.org/10.1177/1077558707305416

39. Escriba-Aguir V, Rodriguez-Gomez M, Ruiz-Perez I. Effectiveness of patient-targeted interventions to promote cancer screening among ethnic minorities: A systematic review. Cancer Epidemiology 2016;44:22–39. doi: http://dx.doi.org/10.1016/j.canep.2016.07.009

40. Glick S, Clarke A, Blanchard A, et al. Interventions to improve minority health and reduce racial and ethnic disparities in care for cervical cancer: A systematic review. Journal of General Internal Medicine 2012;2):S233.

41. Hankerson SH, Weissman MM. Church-based health programs for mental disorders among african americans: A review. Psychiatric Services 2012;63(3):243–49. doi: http://dx.doi.org/10.1176/appi.ps.201100216

42. Lee-Tauler SY, Eun J, Corbett D, et al. A systematic review of interventions to improve initiation of mental health care among racial-ethnic minority groups. Psychiatric Services 2018;69(6):628–47. doi: http://dx.doi.org/10.1176/appi.ps.201700382

43. Jones T, Luth EA, Lin SY, et al. Advance Care Planning, Palliative Care, and End-of-life Care Interventions for Racial and Ethnic Underrepresented Groups: A Systematic Review. Journal of Pain and Symptom Management2021;62(3):e248–e60. doi: http://dx.doi.org/10.1016/j.jpainsymman.2021.04.025

44. Schill K, Caxaj S. Cultural safety strategies for rural Indigenous palliative care: A scoping review. BMC Palliative Care 2019;18(1) (no pagination) doi: http://dx.doi.org/10.1186/s12904-019-0404-y

45. Loutfy M, Tharao W, Logie C, et al. Systematic review of stigma reducing interventions for African/Black diasporic women. Journal of the International AIDS Society 2015;18(1) doi: http://dx.doi.org/10.7448/IAS.18.1.19835

46. Marshall S, Taki S, Laird Y, et al. Cultural adaptations of obesity-related behavioral prevention interventions in early childhood: A systematic review. Obesity Reviews:14. doi: 10.1111/obr.13402

47. Peek ME, Cargill A, Huang ES. Diabetes health disparities: A systematic review of health care interventions. Medical Care Research and Review 2007;64(5 SUPPL.):101S–56S. doi: http://dx.doi.org/10.1177/1077558707305409

48. Fisher TL, Burnet DL, Huang ES, et al. Cultural leverage: interventions using culture to narrow racial disparities in health care. Medical Care Research & Review 2007;64(5 Suppl):243S–82S.

49. Hassen N, Lofters A, Michael S, et al. Implementing anti-racism interventions in healthcare settings: A scoping review. International Journal of Environmental Research and Public Health 2021;18(6):1–15. doi: https://dx.doi.org/10.3390/ijerph18062993

50. McPheeters ML, Kripalani S, Peterson NB, et al. Closing the quality gap: revisiting the state of the science (vol. 3: quality improvement interventions to address health disparities). Evidence report/technology assessment 2012(208.3):1–475.

51. A. Q, M. ON, S. S, et al. Interventions to Improve Minority Health Care and Reduce Racial and Ethnic Disparities. In: Center. E-bSPE, ed.: Department of Veterans Affairs Veterans Health Administration., 2011.

52. Truong M, Paradies Y, Priest N. Interventions to improve cultural competency in healthcare: a systematic review of reviews. BMC health services research 2014;14:99. doi: http://dx.doi.org/10.1186/1472-6963-14-99

53. Anderson LM, Adeney KL, Shinn C, et al. Community coalition-driven interventions to reduce health disparities among racial and ethnic minority populations. Cochrane Database of Systematic Reviews 2015;2015(6) (no pagination) doi: http://dx.doi.org/10.1002/14651858.CD009905.pub2

54. Paluck EL, Porat R, Clark CS, et al. Prejudice Reduction: Progress and Challenges. Annual review of psychology 2021;72:533–60. doi: http://dx.doi.org/10.1146/annurev-psych-071620-030619

55. Davis AM, Vinci LM, Okwuosa TM, et al. Cardiovascular health disparities: A systematic review of health care interventions. Medical Care Research and Review 2007;64(5 SUPPL.):29S–100S. doi: http://dx.doi.org/10.1177/1077558707305416

56. World Health Organisation. Participation as a driver of health equity. Copenhagen.: WHO Regional office for Europe., 2019.

57. M. M. Fair Society, Healthy Lives: The Marmot Review. London: Strategic Review of Health Inequalitites in England post-2010., 2010.

58. White M, Adams J, Heywood P. How and why do interventions that increase health overall widen inequalities within populations? In: Babones S, ed. Health, inequality and society. Bristil: Policy Press 2009.

59. Hassen N, Lofters A, Michael S, et al. ACCEPT Implementing anti-racism interventions in healthcare settings: A scoping review. International Journal of Environmental Research and Public Health 2021;18(6):1–15. doi: https://dx.doi.org/10.3390/ijerph18062993

60. England N. London Workforce Race Strategy London, 2020.

