## Appendix 1 for "A scoping umbrella review to identify anti-racist interventions to reduce ethnic disparities in health and care"

**Appendix 1: search terms and search strategy**

Searcher: Jenni Ford

Person requesting search: Leah DeSouza-Thomas, Sophie Carter

Date of request: 05/01/2022	Date results sent: 18/01/2022

Level of search: (2) annotated bibliography

### Search question:

What works in antiracism

### Terms used:

(racism or racist*) OR (antiracism or anti-racism or antiracist or anti-racist) OR ((race or racial) adj3 (bias* or discriminat* or inequalit* or inequit* or equalit* or disparit*))

AND

intervention* OR program* OR affirmative action OR (policy or policies) OR (strategy or strategies) OR ((education* or training) adj3 (program* or intervention*))

The complete search strategy is below.

### Limits applied:

| **Age group** | **Language** | **Publication type** | **Time limit** |
| --- | --- | --- | --- |
|  | English | Reviews | 2000-current |

### List of databases searched:

**Database**

Embase

Medline

Social Policy and Practice

Social Care Online

Web of Science

### Search strategy

Database: Embase <1974 to 2022 January 11>

Search Strategy:

--------------------------------------------------------------------------------

1	racism/ (9935)

2	(racism or racist*).ab. /freq=2 or (racism or racist*).ti,kw. (3867)

3	(antiracism or anti-racism or antiracist or anti-racist).tw,kw. (510)

4	((race or racial) adj3 (bias* or discriminat* or inequalit* or inequit* or equalit* or disparit*)).tw. (20836)

5	or/1-4 (29058)

6	intervention*.ti,kw. (250539)

7	exp *policy/ (97522)

8	program*.ti,kw. (269293)

9	affirmative action.tw. (425)

10	(policy or policies).ti,kw. (71326)

11	(strategy or strategies).ti,kw. (198658)

12	social justice/ and (policy or policies or strateg* or program* or intervention*).tw. (2694)

13	exp *education/ and (intervention* or program*).tw. (172785)

14	((education* or training) adj3 (program* or intervention*)).tw. (179586)

15	or/6-14 (1034207)

16	5 and 15 (2030)

17	limit 16 to (english language and yr="2000 -Current") (1968)

18	limit 17 to "reviews (best balance of sensitivity and specificity)" (232)

***************************

Database: Ovid MEDLINE(R) ALL <1946 to January 11, 2022>

Search Strategy:

--------------------------------------------------------------------------------

1	*Racism/ (3659)

2	(racism or racist*).ab. /freq=2 or (racism or racist*).ti,kw. (3665)

3	(antiracism or anti-racism or antiracist or anti-racist).tw,kw. (497)

4	((race or racial) adj3 (bias* or discriminat* or inequalit* or inequit* or equalit* or disparit*)).tw. (15313)

5	or/1-4 (19736)

6	intervention*.ti,kw. (185235)

7	exp Public Policy/ (148223)

8	program*.ti,kw. (228707)

9	affirmative action.tw. (419)

10	(policy or policies).ti,kw. (66455)

11	(strategy or strategies).ti,kw. (160991)

12	Social Justice/lj, st (555)

13	exp *Education/ and (intervention* or program*).tw. (154551)

14	((education* or training) adj3 (program* or intervention*)).tw. (134275)

15	or/6-14 (887287)

16	5 and 15 (1538)

17	limit 16 to (english language and yr="2000 -Current") (1454)

18	limit 17 to "reviews (best balance of sensitivity and specificity)" (191)

***************************

Database: Social Policy and Practice <202110>

Search Strategy:

--------------------------------------------------------------------------------

1	(racism or racist*).ab. /freq=2 or (racism or racist*).ti. (773)

2	(antiracism or anti-racism or antiracist or anti-racist).tw. (391)

3	((race or racial) adj3 (bias* or discriminat* or inequalit* or inequit* or equalit* or disparit*)).tw. (1499)

4	intervention*.ti. (5666)

5	program*.ti. (7399)

6	affirmative action.tw. (40)

7	(policy or policies).ti. (9868)

8	(strategy or strategies).ti. (4798)

9	((education* or training) adj3 (program* or intervention*)).tw. (4499)

10	1 or 2 or 3 (2321)

11	4 or 5 or 6 or 7 or 8 or 9 (29869)

12	10 and 11 (192)

13	limit 12 to yr="2000 -Current" (122)

***************************
