## Appendix 2 for "A scoping umbrella review to identify anti-racist interventions to reduce ethnic disparities in health and care"

Appendix 2: AMSTAR2 Critical and Non-critical items

| Critical items | Non-critical items |
| --- | --- |
| Protocol registered before commencement of review | PICO used for research question/inclusion criteria |
| Adequacy of the literature search | Justification of study design inclusion |
| Risk of bias from individual studies being included in the review | Study selection completed in duplicate |
| Appropriateness of meta-analytical methods | Data extraction completed in duplicate |
| Consideration of risk of bias when interpreting the results of the  review | Justification for excluding individual studies |
| Assessment of presence and likely impact of publication bias | Adequate description of included studies |
|  | Sources of funding of included studies |
|  | Impact of risk of bias on meta-analysis or other evidence synthesis  assessed |
|  | Satisfactory explanation and discussion of heterogeneity of results |
|  | Conflicts of interest of authors declared |
