## Appendix 3 for "A scoping umbrella review to identify anti-racist interventions to reduce ethnic disparities in health and care"

| **Appendix 3: AMSTAR2 Quality rating of included reviews** | | | | | | | | | | | | | | | | | |
| --- | --- | --- | --- | --- | --- | --- | --- | --- | --- | --- | --- | --- | --- | --- | --- | --- | --- |
|  | AMSTAR2 rating | | | | | | | | | | | | | | | | |
| **Full Citation** | Q1 | Q2 | Q3 | Q4 | Q5 | Q6 | Q7 | Q8 | Q9 | Q10 | Q11 | Q12 | Q13 | Q14 | Q15 | Q16 | Overall rating |
| Davis AM, Vinci LM, Okwuosa TM, Chase AR, Huang ES. Cardiovascular Health Disparities. Medical Care Research and Review. 2007;64(5_suppl):29S-100S. | N | N | N | Y | Y | Y | N | N | N | N | N/A | N/A | N | N | N/A | N | Critically low |
| Engberg, M. E. (2004). "Improving intergroup relations in higher education: A critical examination of the influence of educational interventions on racial bias." Review of Educational Research 74(4): 473-524. | N | N | N | N | N | N | N | N | N | N | N/A | N/A | N | N | N/A | N | Critically low |
| Escriba-Aguir, V., et al. (2016). "Effectiveness of patient-targeted interventions to promote cancer screening among ethnic minorities: A systematic review." Cancer Epidemiology 44: 22-39. | Y | N | N | Y | Y | Y | N | N | N | N | N/A | N/A | N | N | N/A | Y | Critically low |
| Fisher T. L., D. L. Burnet, E. S. Huang, M. H. Chin and K. A. Cagney. Cultural leverage: interventions using culture to narrow racial disparities in health care. Medical Care Research & Review 2007 Vol. 64 Issue 5 Suppl Pages 243S-82S. | N | N | Y | Y | Y | Y | N | N | N | N | N/A | N/A | N | Y | N/A | Y | Critically low |
| Glick, S., Clarke, A. R., Blanchard, A., et al. 2012. Interventions to improve minority health and reduce racial and ethnic disparities in care for cervical cancer: A systematic review. Journal of General Internal Medicine 2) S233. | N | N | N | Y | N | N | N | Y | Y | N | N/A | N/A | Y | N | N/A | Y | Critically low |
| Hankerson S.H. and M. M. Weissman. Church-based health programs for mental disorders among african americans: A review. Psychiatric Services 2012 Vol. 63(3) Pages 243-249 | N | N | N | Y | N | N | N | N | N | N | N/A | N/A | N | N | N/A | Y | Critically low |
| Hassen N., A. Lofters, S. Michael, A. Mall, A. D. Pinto and J. Rackal. Implementing anti-racism interventions in healthcare settings: A scoping review. International Journal of Environmental Research and Public Health 2021 Vol. 18(6) Pages 1-15 | N | N | N | Y | Y | Y | N | Y | N | N | N/A | N/A | N | Y | N/A | Y | Critically low |
| Schill K. and S. Caxaj. Cultural safety strategies for rural Indigenous palliative care: A scoping review. BMC Palliative Care 2019 Vol. 18(1) (no pagination) | N | N | N | Y | N | Y | N | N | N | N | N/A | N/A | N | N | N/A | Y | Critically low |
| Truong M., Y. Paradies and N. Priest. Interventions to improve cultural competency in healthcare: a systematic review of reviews. BMC health services research 2014 Vol. 14 Pages 99 | N | N | N | Y | Y | Y | N | N | N | N | N/A | N/A | N | Y | N | Y | Critically low |
| Waller. Broken fixes: A systematic analysis of the effectiveness of modern and postmodern interventions utilized to decrease IPV perpetration among Black males remanded to treatment. Aggression and Violent Behavior 2016 Vol. 27 Pages 42-49. Accession Numb | N | N | N | N | N | N | N | N | N | N | N/A | N/A | N | N | Y | N | Critically low |
| Anderson LM, Adeney KL, Shinn C, Safranek S, Buckner‐Brown J, Krause LK. Community coalition‐driven interventions to reduce health disparities among racial and ethnic minority populations. Cochrane Database of Systematic Reviews 2015, Issue 6. Art. No.: CD009905. DOI: 10.1002/14651858.CD009905.pub2. Accessed 10 January 2023. | Y | Y | Y | Y | Y | Y | Y | Y | Y | N | Y | Y | Y | Y | Y | Y | High |
| Jones T., E. A. Luth, S. Y. Lin and A. A. Brody. Advance Care Planning, Palliative Care, and End-of-life Care Interventions for Racial and Ethnic Underrepresented Groups: A Systematic Review. Journal of Pain and Symptom Management 2021 Vol. 62(3) Pages e248-e260 | Y | Y | Y | Y | Y | Y | N | Y | Y | N | N/A | N/A | Y | Y | N/A | Y | Low |
| Lee-Tauler S.Y., J. Eun, D. Corbett and P. Y. Collins. A systematic review of interventions to improve initiation of mental health care among racial-ethnic minority groups. Psychiatric Services 2018 Vol. 69(6) Pages 628-647 | Y | Y | N | Y | Y | Y | N | Y | Y | N | N/A | N/A | Y | N | N/A | Y | Low |
| Loutfy M., W. Tharao, C. Logie, M. A. Aden, L. A. Chambers, W. Wu, et al.Systematic review of stigma reducing interventions for African/Black diasporic women. Journal of the International AIDS Society 2015 Vol. 18 Issue 1 | Y | Y | N | Y | Y | Y | Y | Y | Y | N | N/A | N/A | Y | N/A | N/A | Y | Low |
| Marshall, S, Taki, S, Laird, Y, Love, P, Wen, LM, Rissel, C. Cultural adaptations of obesity-related behavioral prevention interventions in early childhood: A systematic review. Obesity Reviews. 2022; 23( 4):e13402. doi:10.1111/obr.13402 | N | Y | N | Y | Y | Y | N | Y | Y | N | N/A | N/A | Y | Y | N/A | Y | Low |
| McPheeters M.L., S. Kripalani, N. B. Peterson, R. T. Idowu, R. N. Jerome, S. A. Potter, et al. Closing the quality gap: revisiting the state of the science (vol. 3: quality improvement interventions to address health disparities). Evidence report/technology assessment 2012 Issue 208 .3 Pages 1-475 | Y | N | Y | Y | Y | Y | Y | Y | Y | N | N/A | N/A | N | N | N/A | Y | Low |
| Paluck E.L., R. Porat, C. S. Clark and D. P. Green. Prejudice Reduction: Progress and Challenges. Annual review of psychology 2021 Vol. 72 Pages 533-560 | N | Y | Y | Y | Y | Y | N | Y | Y | N | Y | Y | Y | Y | Y | Y | Low |
| Peek ME, Cargill A, Huang ES. Diabetes Health Disparities. Medical Care Research and Review. 2007;64(5_suppl):101S-156S. doi:10.1177/1077558707305409 | Y | Y | N | Y | Y | Y | N | Y | Y | N | N/A | N | N | N | N/A | Y | Low |
