## Appendix 4 for "A scoping umbrella review to identify anti-racist interventions to reduce ethnic disparities in health and care"

#### Appendix 4. Characteristics of interventions

Interventions targeting the individual and community level of the socioecological model were more common amongst the studies included in our review.

#### Individual and community level interventions

Individual and community level interventions aimed to change knowledge, attitudes, behaviour on health and healthcare, and develop community partnerships. Interventions can be grouped into four categories: education, access, use of community health workers and partnership-building. Education interventions were most identified amongst interventions targeting individuals and are described according to content and delivery.

##### Education

Patient education interventions for common chronic conditions targeted knowledge and behaviours around risk factors for cardiovascular disease and diabetes like diet, exercise and stress (^36 45 51^, or self-management of asthma ^46^ or diabetes ^45 48 51^. One review which focused specifically on community coalition-driven interventions (characterised by representation from multiple community sectors who collaborate to implement bottom-up decision-making and use of local assets and resources to build community capacity ^59^) ^51^. Health education was identified as part of broad-scale community system-level change and one of four core community engagement interventions identified in the review. Coalition-driven education interventions aim to create sociocultural and physical environments which support healthy choices and improve quality of life e.g. community-based adult nutrition classes and school-based nutrition education for children ^51^.

Church-based mental health education included health promotion programmes to increase knowledge of causes, treatment and sources of help for anxiety and depressive symptoms, and health promotion programmes to change risky behaviours in substance-related disorders ^39^. Health system-based mental health education aimed to educate patients about psychotherapy entry and attendance, improve depression or psychosis literacy, reduce stigma associated with antidepressants, and increase access to help-seeking ^40^.

HIV interventions targeting African American women included reframing of traumatic events and reducing negative thoughts, enhancing HIV-knowledge, building psychological skills for young people recently diagnosed with HIV, and a maternal HIV self-management intervention designed to improve HIV-related knowledge, reduce emotional distress and promote care-seeking strategies among low-income African American mothers with HIV ^43^. Participatory educational exercises addressed misinformation around HIV, raised awareness of HIV-related stigma and developed skills to addressing stigmatizing situations ^43^.

Cancer education targeted knowledge of breast self-exam, and colorectal, breast and cervical cancer symptoms ^37 38 51^.

Patient education for palliative care focused on advanced care planning. Interventions were designed to improve utilisation of end-of-life services for people from ethnic minority groups by addressing barriers including language, knowledge, attitudes and beliefs about end-of-life services, and improving health literacy ^41^. Examples included using video stories, narratives, and testimonials to model how to engage in advanced care planning ^46^.

##### Facilitating access

Interventions focused on access were identified for cancer detection and treatment. These included free colorectal cancer tests, counselling, test reminders, small media for colorectal, breast and cervical cancer ^37^. Interventions to increase access to cervical cancer screening included scheduling appointments, low-cost sources of care and transportation and telephone counselling and support ^38^. Interventions to improve diagnosis and treatment of premalignant disease included telephone counselling, a streamlined process for follow-up of abnormalities, intensive follow-up and/or vouchers for reduced cost care, a personalized letter, pamphlet, audio-visual presentation or transportation incentives, and messages in the media ^38^.

##### Community partnership-building

Partnership-building was identified as a community level intervention with regard Aboriginal and Indigenous communities ^42 47^. This involved actively engaging Indigenous communities at multiple levels in healthcare planning and throughout the process ^47^. Partnership-building also involved reorganising power by strengthening cohesion within communities, forming caucus groups and encouraging anti-racist community organizing ^47^.

The review of cultural safety strategies in rural Indigenous palliative care also emphasized the importance of community ownership of palliative services ^42^. The review suggests that this can be achieved by ensuring the needs of communities are met and building on community strengths and assets. For example, partnering with community organizations or Indigenous tribe leaders in programme development, delivery and evaluation, or engaging a community advisory committee in service planning and delivery with the aim of building community ownership of services over time ^42^.

##### Cultural adaptation

A common theme amongst the individual level interventions was cultural tailoring and adaptation. Two reviews focused on how interventions incorporated cultural adaptations and cultural safety strategies.

The first is a review looking at cultural adaptation of obesity-related behaviour prevention interventions in children aged 0-5 (those targeting feeding, nutrition, activity and/or sleep) and excluded interventions that involved language translation only ^44^. The review uses cultural sensitivity theory ^60^ to categorise cultural adaptations into “surface” and “deep” structures where surface structures involved matching intervention materials and messages to observable ("superficial") characteristics of the target population, such as modifying language and translations, and altering activities to improve suitability ^44 60^. These were most commonly seen. Deep structures involve “incorporating the cultural, social, historical, environmental and psychological forces that influence the target health behaviour in the proposed target population” ^60^. These were less commonly seen but included addressing cultural values in the intervention content (“deep structure”) and involving culturally-matched intervention facilitators (“surface” and “deep” structures) ^44^.

The second is the review of cultural safety strategies for palliative care in rural Indigenous communities in Canada, the United States, New Zealand and Australia. The review revealed a number of interventions incorporating the cultural and social forces in these populations. These included symbolic or small gestures that welcome Indigenous palliative clients such as displaying Indigenous art, acknowledging National Aboriginal Day, integrating culturally appropriate food in menus and respecting spiritual practices even in the absence of spiritual care services (Schill). Other interventions involved anticipating barriers to care, shared decision-making that includes active patient and family involvement, respectful, clear, and culturally appropriate communication, and empowering cultural identity by acknowledging culturally-specific beliefs and traditions as legitimate during the palliative care process and are essential to decision-making ^42^.

The cultural adaptations described by the remaining reviews are grouped according to whether surface (matching materials or messages to observable characteristics) or deep cultural adaptations (incorporating cultural, social, historical, environmental and psychological forces into interventions) were used.

###### Surface cultural adaptations

In terms of materials, culturally adapted end-of-life care involved providing bilingual materials and using creative and diverse approaches including video soap opera (telenovela) targeting the Hispanic community in the US ^39^.

###### Deep cultural adaptations

Deep cultural adaptations for mental health programmes targeting African Americans involved emphasizing black culture and spirituality, including using Churches as a setting for the intervention, involving Church mentors, distributing gospel music and Biblical scriptures, and including prayer ^39^.

##### Lay Community Health Workers

Lay Community Health Workers (CHWs) were identified in reviews of interventions for asthma, diabetes, HIV/AIDS prevention and pesticide exposure in migrant farm workers ^51^, cancer screening ^46^, smoking cessation ^51^, and cardiovascular disease ^36 46^. CHWs provided enhanced case tracking and follow-up, home visits for education and supplies, and community health fairs. CHWs either acted alone or in collaboration with a nurse, and findings from included studies suggest they were relatively more successful than physician-led services ^36^. One review described CHWs who came from the same community as the target population as among the most successful strategies emerging from the findings. This was put down to their understanding the culture of the community, and ability to develop relationships with patients which overcome common obstacles on this basis ^46^. Despite the several reviews identifying CHWs as a potentially effective intervention, there was a lack of description of CHWs characteristics, recruitment or training.

#### Healthcare organisation level interventions

Healthcare organisation level interventions can be grouped into three categories: organisation of care, clinician interactions with patients, and workforce and leadership. Within organisation of care interventions included collaborative care, case management and colocation of services. Interventions involving clinician interactions with patients included practice guidelines, decision support tools and computerised reminders. Workforce and leadership interventions included training in diversity and cultural competency, commitment from those in leadership, anti-racism quality improvement initiatives and workforce recruitment and retention policies.

##### Organisation of care

###### Collaborative care

A review of interventions to improve initiation of mental health care among racial-ethnic minority groups found that more than one-third of included studies involved collaborative care interventions, and a larger body of evidence supporting collaborative care than any other intervention ^40^. Several components of collaborative care were identified from included studies appear to be effective, including a case manager supporting primary care providers with patient evaluation, education, medication, psychosocial treatment and linkage to mental health services ^40^. Another approach was for a cancer-depression clinical specialist to provide psychotherapy and care management in oncology clinics ^40^.

A review of quality improvement interventions to address health disparities found that evidence from a number of studies suggests that collaborative care is worthy of future study and possibly wider dissemination ^48^. The literature emphasized the need for coordination of care by multiple clinical providers, usually by a care manager ^48^.

###### Case management

The review of interventions to improve initiation of mental health care among racial-ethnic minority groups identified case management strategies involving a multidisciplinary team approach, frequent contacts with patients, small caseloads and assertive outreach. There was evidence that the strategies resulted in significant improvement in the receipt of psychiatric care but initiation of care didn’t differ by ethnicity ^40^.

###### Colocation of services

The review of interventions to improve initiation of mental health care among racial-ethnic minority groups also found that colocation of mental health services (provision of mental health services in primary care clinics) and providing telepsychiatry at a community health centre improved initiation of care and antidepressant use in some groups ^40^. The review suggested that the practical convenience and privacy of seeking care in primary care settings may enhance help-seeking in ethnic minority groups ^40^.

##### Clinician interactions with patients

A review of diabetes health disparities found a number of studies targeting clinician behaviour, the majority of which involved the application of generic diabetes quality improvement initiatives to ethnic minority groups ^45^. The interventions typically included practice guidelines, continuing medical education, computerized decision- support reminders, in-person feedback and problem-based learning. The interventions resulted in improved processes of care (HbA1c monitoring, foot care, exercise counselling etc) and improved diabetes control. None of the interventions included culturally tailored components ^45^. However improved care and control is particularly relevant to ethnic minorities as evidence suggests they are less likely to have access to care and more likely to have worse control of their diabetes ^45^. This suggests **that targeting clinicians for quality improvement in service delivery in areas with higher proportions of ethnic minority populations may be an effective strategy** for improving diabetes outcomes in these groups ^45^.

##### Workforce and leadership

###### Diversity training

A review of the implementation of anti-racism interventions in healthcare settings identified several articles focused on developing tools, training, workshops or curricula ^47^. Key factors for effective race equity training identified from the literature include whether the healthcare organization has a formal anti-racism policy, whether race equity training is voluntary or mandatory, cost of attendance, facilitator qualifications, impact evaluation, whether training is ongoing rather than a “one-off”, and whether training sufficiently addresses healthcare provider-specific issues ^47^. The review also found that anti-racism work to support systemic change with healthcare institutions should be linked to broader concepts of power, hierarchy and dominance, and be informed by critical theories ^47^. A review of interventions to improve cultural competency in healthcare found a number of studies of cultural competency training and workshops for health practitioners (doctors, nurses and community health workers) ^50^.

###### Leadership

The review of the implementation of anti-racism interventions in healthcare settings also identified that strong, consistent visible leadership that addresses racism is important for sustainable organizational change ^47^. The literature suggests that leaders and governing bodies at different levels of organization need to be involved in anti-racism initiatives, with executive teams completing training first and providing support to staff.

###### Recruitment and retention policies

One review identified that organizational and human resource policies in healthcare organizations can contribute to racial health disparities, and that explicit anti-racist policies should be developed ^47^.

#### Interventions outside of the healthcare setting

###### Education

A review of the influence of education interventions on racial bias in higher education adds to the findings on diversity training and workshops ^32^. A number of education interventions were identified including multicultural course interventions (e.g. mandatory/ non-mandatory diversity courses; ethnic studies and women's studies), diversity workshop and training interventions, peer-facilitated interventions (e.g. peer training, learning communities, collaborations, intergroup dialogue), and community service interventions (e.g. volunteer work) ^32^.

A review or prejudice reduction interventions looked at interventions aimed at college and grade or high school students ^52^. The review identified a number of educational interventions including cognitive and emotional training, social categorization, peer influence and dialogue, value consistency and self-worth training, interpersonal contact, multicultural antibias moral education, diversity training, cooperative learning, intercultural training and conflict resolution ^52^.

###### Criminal justice

One review was identified which was set in the criminal justice system ^34^. The review looked at interventions to decrease intimate partner violence (IPV) perpetration among Black males remanded to treatment. The review compared interventions based in modern therapy approaches such as cognitive behavioural therapy (CBT) and psychoeducation, with postmodern therapy approaches using goal-setting interventions which are strengths-based and developed to empower marginalized and disenfranchised populations who may be less educated, underemployed and stigmatized. It allows a shift in power to the client, allowing him to develop treatment goals and collaborate with the therapist.^34^
